# Zero-COVID Policies: Melbourne’s 112-Day Hard Lockdown Experiment Harmed Mostly Mothers

**DOI:** 10.1101/2022.01.30.22270130

**Authors:** Stefanie Schurer, Kadir Atalay, Nick Glozier, Esperanza Vera-Toscano, Mark Wooden

## Abstract

Lockdowns were used worldwide to mitigate the spread of SARS-CoV-2. We demonstrate that the 112-day hard lockdown in Melbourne, Australia, the longest among OECD jurisdictions, exclusively penalized families with young children. To identify the causal impact of lockdown, we interrogated nationally-representative longitudinal survey data and exploited quasi-experimental variation in Melbourne’s lockdown, one that left other jurisdictions unaffected. Using difference-in-differences estimation, we found that, surprisingly, most vulnerable groups (the young, poor, lonely and those with previous mental health conditions) were left unscathed. However, we found mothers experienced significant, sizable declines in health and work hours, and increases in loneliness, despite feeling safer and being more active. Zero-COVID policies are not as harmful as may have been expected but came at high cost to mothers in society.

**One-Sentence Summary:** Melbourne’s hard lockdown left most vulnerable groups unscathed but led to greater ill-health and loneliness in mothers.

## 1. Introduction

Of all the non-pharmaceutical interventions (NPI) introduced to mitigate the spread of SARS-CoV-2 (COVID-19), none has been so ‘intrusive’ and ‘drastic’ as the lockdown (*1*). It is commonly understood as a restriction of the individual right to move freely which ‘applies to all people’, albeit some categorization systems label such mobility restrictions as ‘partial lockdown’, ‘household confinement’, or ‘movements for non-essential activities forbidden’ (*2*). By the end of April 2020, half of humanity was in some form of lockdown: Almost 4 billion people across 90 countries were asked by their governments to stay at home (*3*). By the end of 2021, some countries had ordered their residents into their eighth lockdown, while other places had experienced over 100 days sheltered in place.

Early on in the pandemic, stay-at-home orders with all their obvious personal costs, were accepted as an essential strategy to set up pandemic response systems and to prevent health services from being overwhelmed (*4–5*). During the second wave in 2020, however, an international scientific debate erupted (*6*) over whether community spread could only be controlled through maximum suppression (elimination), which would have required hard lockdowns and border closures (Zero-COVID strategy) (*7–8*), or whether less restrictive mitigation measures would achieve the same goal (*9*). Evidence exists both in favor of elimination strategies to achieve their primary objective (*10–13*) and against them (*14–16*), while some proposed that less intrusive NPI may be more appropriate should infection numbers surge (*1*).

However, Zero-COVID strategies cannot be used indefinitely, as lockdown requires many sacrifices from residents (*17*). Such sacrifices include poorer mental health (*18–19*), increases in loneliness because of the social isolation (*20–22*), and the abandonment of healthy or adoption of unhealthy behaviours to cope with that isolation. Importantly, a great concern was that lockdowns exacerbated social, economic and gender inequalities (*23–27*), harming in particular families (*28*). The burden of lockdown to mothers was at the forefront of the popular debate, with some descriptive studies showing that the wellbeing of mothers was likely to be most impaired (*29–30*). The closure of schools or child-care centres meant children had to be cared for and taught at home. Women were expected to cover such home-schooling or -caring duties (*31–33*). The impact of both the pandemic and lockdowns on mothers’ labor supply were deemed substantial (*32, 34–36*).

Although many studies exist on the collateral damage of lockdowns and their unequal impact, most rely on before-after comparisons (e.g. *18,28, 33*), failing to identify lockdown’s causal impact. The reason is that lockdowns are not imposed exogenously. First, in many jurisdictions, such as the UK, the USA, and many European countries, lockdown was a last-resort policy measure, that occurred after a string of events, with the aim of flattening the curve of exponentially growing caseloads, patients in ICUs and infected humans dying from COVID-Thus, lockdown costs may be severely overestimated, as they are overshadowed by COVID-related trends of increasing morbidity and mortality. Second, lockdowns caused severe economic contraction, job loss, and business closures at the macroeconomic level. Thus, lockdown effects are potentially confounded by income shocks that threaten the financial sustainability of households. Third, lockdowns were implemented some time after lockdown announcements, which may have already changed behaviors pre-lockdown. This could lead to a severe underestimate of the effect of lockdown.

In this paper, we describe how we used methods from the economic policy evaluation literature (difference-in-difference estimation models) in combination with high-quality, nationally representative longitudinal survey data to overcome these identification issues. We studied a unique natural experiment that arose from Australia’s second 2020 lockdown (Fig. 1). On 9 July 2020, Australia’s second largest city, Melbourne, which is also the capital of the state of Victoria, was locked down for 112 consecutive days, using an aggressive Zero-COVID strategy to return to zero infections (*37–38*), while other Australian jurisdictions remained open. The Melbourne case is of paramount scientific importance. Melbourne endured the second-longest consecutive hard lockdown in 2020 (after Buenos Aires) and holds the unenviable record of greatest cumulative time spent in lockdown worldwide. Melbourne’s Zero-COVID achievement after this hard and long lockdown was hailed by the scientific community (*39*).

**Figure 1.**
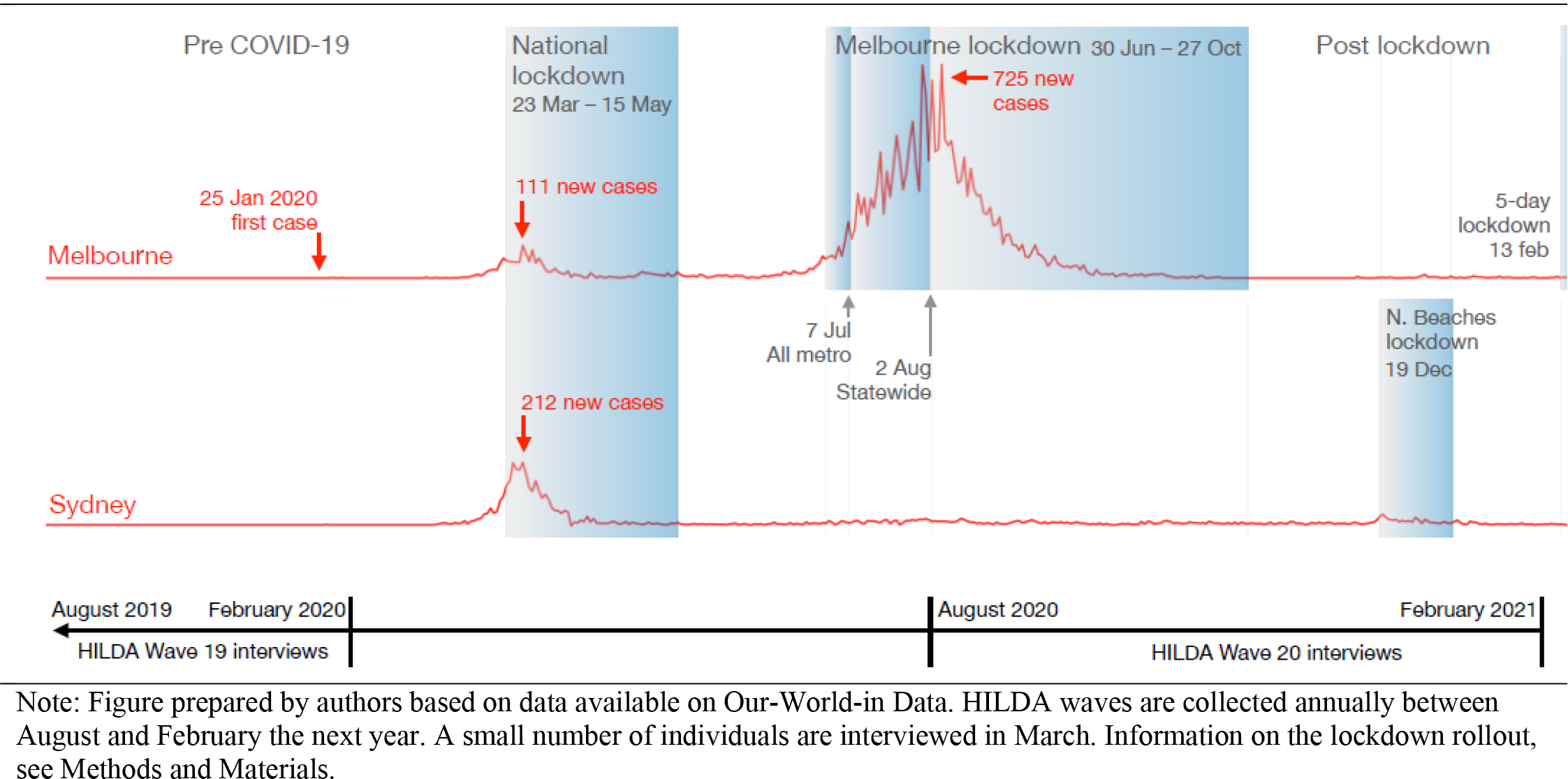
Description of the natural experiment with a timeline of COVID-19 infection numbers, lockdown dates, and HILDA data collection windows

Thanks to this natural experiment, which provided a valid control group outside of Melbourne/Victoria, our study was the first to rigorously evaluate the causal impact of lockdown on human life across multiple domains (health, health behaviours, social connectedness, labor supply and income). In the analysis, we highlighted the inequalities of lockdown penalties. Melbourne’s hard and long lockdown unequivocally penalized mothers’ health, social connectedness, and labor supply, while it left fathers and other potentially vulnerable groups (the resource poor, the lonely, and those with high health care needs) relatively unscathed.

## 2. The lockdown natural experiment

Melbourne’s second lockdown occurred after a successful national lockdown (23 March-15 May 2020), when life in Australia seemed to be returning to normality. New daily infections numbered less than 20, school students had returned to the classroom, workers were permitted to return to the office, and service industries were allowed to operate again, albeit with capacity limits. In late June, breaches in the hotel quarantine system saw case numbers in Melbourne begin to rise again. This led to the reimposition of stay-at-home orders (Stage 3 restrictions) in certain postcodes in the western and northern suburbs of Melbourne on 2 July before being extended to all areas of Melbourne, as well as one shire on the outskirts of Melbourne, on 9 July. Borders closed between New South Wales, the most populous state in Australia, and Victoria on the same day. On 2 August the Victorian Government declared a state of disaster and imposed an 8 pm to 5 am curfew in Melbourne while regional Victoria moved to less severe restrictions. Lockdown restrictions were lifted in regional Victoria on 17 September and the curfew in Melbourne was lifted on 28 September. Stay-at-home orders in Melbourne were removed on 28 October.

The Melbourne lockdown provided an almost perfect policy laboratory. With international and state borders closed, Melbournian residents could not easily escape lockdown measures. Compliance with lockdown orders was relatively high in Melbourne and relatively low in the rest of Victoria, which is one reason why we studied the Melbourne case only (see Materials and Methods). In addition, the lockdown occurred in the absence of widespread financial difficulty, due to the availability of JobKeeper, a generous federal pay-replacement program, which buffered businesses’ profit loss, while keeping workers employed with compensated pay, and a substantial supplement to the incomes of those relying on income-support payments. Also, in contrast to the international experience, the Melbourne lockdown commenced when daily new infections were less than 100. Even during the lockdown, the maximum number of new daily infections peaked at 725 (Fig. 1) and the deaths that did subsequently occur during lockdown were among those aged 70 years or older, with 75% occurring in residential care.

Thus, the cases did not cause a health burden of disease among youth, the working age or younger retiree populations that were most affected in their daily activities by lockdown.

Meanwhile, in the rest of the country, including Sydney, Australia’s largest city and capital of New South Wales, residents could move freely. Most businesses were allowed to operate, including the hospitality sector, even if with some capacity or density limits. The only exception was a five-day mobility restriction in one out of 30 government areas in the Greater Sydney area in mid-December 2020, and even that did not involve a stay-at-home order.

## 3. Data, sample and outcome definition

Fortuitously, during Melbourne’s lockdown period, the 20^th^ wave of Australia’s nationally representative Household, Income, and Labour Dynamics in Australia (HILDA) survey was undertaken (Fig. 1). This household panel commenced in 2001 with a nationally representative sample of Australian households (see Methods and Materials). In line with data-collection protocols in previous years, almost all 2020 HILDA Survey participants (>93%) were interviewed between August and late October, which fell directly within the window of Melbourne’s lockdown experiment. Another 3.4% were interviewed in November, a period that covers the potential spill-over effects of lockdown.

We used these data and knowledge of the exact rollout timing of the Melbourne lockdown to quantify the effect of lockdown on human life. We compared the outcomes in Melbourne (population 5.16 million) with the outcomes of Sydney (population 5.37 million), the most comparable city to Melbourne, which was not locked down. In the HILDA Survey data we had 9,441 individuals (*n*) or 60,108 person-year observations (*pyo*) who either live in Sydney or in Melbourne to tell the tale of two cities located at the extreme ends of available mobility restrictions.

From these data, we derived proxies that capture comprehensively and at the broadest level human life across four domains. For each domain, we identified three measures that have been widely used in previous public health and applied economic research (table S1) — (i) Health (all derived from the SF-36 inventory – see Materials and Methods): mental health, which refers to symptoms of anxiety and depression (0–100), general health (0–100), and bodily pain (0–100); (ii) Health behaviors: Body Mass Index (kg/m^2^), frequency of physical activity (none- daily), and frequency of alcohol consumption (none-daily); (iii) Perceptions of social connectedness: safety (very dissatisfied [0]-very satisfied [10]), loneliness (strongly disagree [1]-strongly agree [7]), and feeling part of the local community (very dissatisfied [0]-very satisfied [10]); (iv) Labor supply and income: work hours (weekly, incl. 0), salary and wages from all jobs (weekly, in Australian Dollars), and government transfers excluding family benefits (weekly, in Australian Dollars). Time trends in these outcomes are reported in (figs. S1 and S2).

The choice of each outcome measure was based on three considerations: (i) it was consistently recorded annually for the past decade; (ii) its measurement period referred to the current time period, making it suitable for attributing its potential change to lockdown; and (iii) it can be assumed continuous so that it can be scaled to the same standardized unit, which facilitates comparisons of magnitude across outcomes. Sample sizes differed for each outcome but ranged between 45,528*pyo* (8,114*n*) and 52,813*pyo* (8,932*n*) (fig. S1). In addition, we analyzed three outcomes collected in a 2020 COVID-19 module (5713*n*), which asked about individuals’ past COVID-19 experience (positive test=1, otherwise=0) and perceptions of risk of both infection and serious illness (both scaled between 0% and 100%).

## 4. Estimation results

### a. Before-During Comparisons

Many dimensions of human life significantly changed for Melbournians in 2020 when compared against the long-term average (2011–2019) (Table 1), in both positive and negative ways. Mental health declined from 72.55 to 68.65 (*P*<0.001), while BMI increased from 26.5 to 27.2 (*P*<0.001), and feelings of loneliness increased, marginally significantly, from 2.6 to 2.7 (*P*=0.057). Other outcomes changed for the better. For instance, the frequency of physical activity increased from 3.6 to 3.8 (*P*<0.001) and satisfaction with feeling safe from 8.2 to 8.3 (*P*=0.002). Unsurprisingly, average workhours declined from 23.9 hours per week to 20.2 hours per week (*P*<0.001), while average weekly government transfers increased by 50% from $56.6 to $84.5 (*P*<0.001). There were no significant changes in weekly income derived from current wages and salaries (*P*=0.392), the frequency of alcohol consumption (*P*=0.668), or perceptions of feeling part of the local community (*P*=0.275).

**Table 1:**
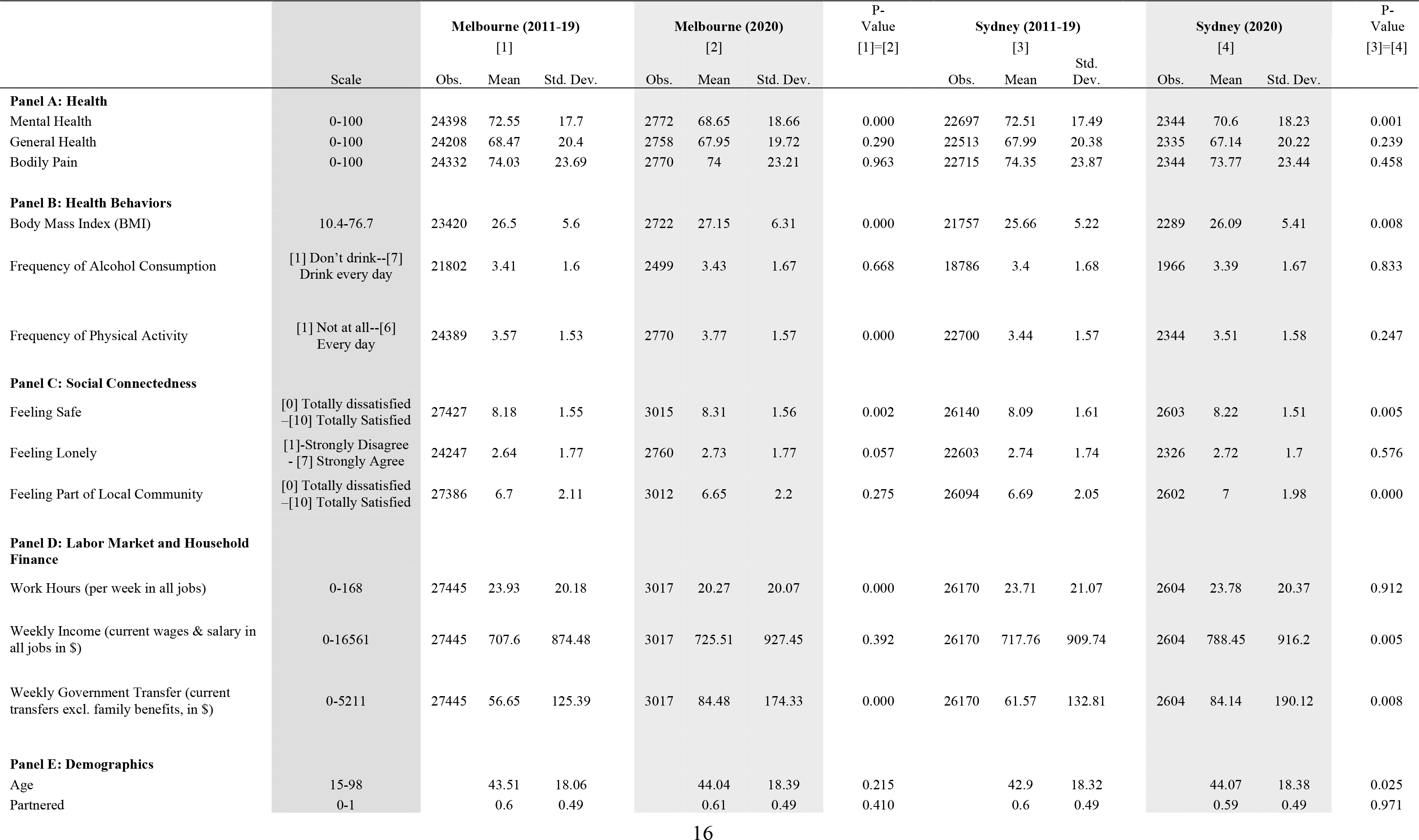
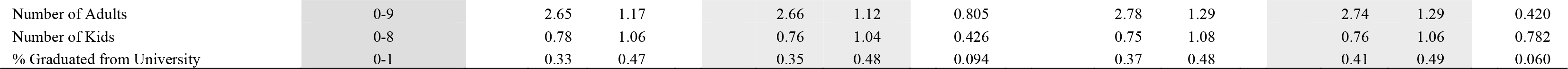
Summary statistics and definitions, separately by treatment (Melbourne) and control (Sydney) group, before and during lockdown

|  |  | Melbourne (2011-19) |  |  | Melbourne (2020) |  |  | P-Value<br>[1]=[2] | Sydney (2011-19) |  |  | Sydney (2020) |  |  | P-Value<br>[3]=[4] |
| --- | --- | --- | --- | --- | --- | --- | --- | --- | --- | --- | --- | --- | --- | --- | --- |
|  | Scale | Obs. | Mean | Std. Dev. | Obs. | Mean | Std. Dev. |  | Obs. | Mean | Std. Dev. | Obs. | Mean | Std. Dev. |  |
| **Panel A: Health** |  |  |  |  |  |  |  |  |  |  |  |  |  |  |  |
| Mental Health | 0-100 | 24398 | 72.55 | 17.7 | 2772 | 68.65 | 18.66 | 0.000 | 22697 | 72.51 | 17.49 | 2344 | 70.6 | 18.23 | 0.001 |
| General Health | 0-100 | 24208 | 68.47 | 20.4 | 2758 | 67.95 | 19.72 | 0.290 | 22513 | 67.99 | 20.38 | 2335 | 67.14 | 20.22 | 0.239 |
| Bodily Pain | 0-100 | 24332 | 74.03 | 23.69 | 2770 | 74 | 23.21 | 0.963 | 22715 | 74.35 | 23.87 | 2344 | 73.77 | 23.44 | 0.458 |
| **Panel B: Health Behaviors** |  |  |  |  |  |  |  |  |  |  |  |  |  |  |  |
| Body Mass Index (BMI) | 10.4-76.7 | 23420 | 26.5 | 5.6 | 2722 | 27.15 | 6.31 | 0.000 | 21757 | 25.66 | 5.22 | 2289 | 26.09 | 5.41 | 0.008 |
| Frequency of Alcohol Consumption | [1] Don't drink--[7] Drink every day | 21802 | 3.41 | 1.6 | 2499 | 3.43 | 1.67 | 0.668 | 18786 | 3.4 | 1.68 | 1966 | 3.39 | 1.67 | 0.833 |
| Frequency of Physical Activity | [1] Not at all--[6] Every day | 24389 | 3.57 | 1.53 | 2770 | 3.77 | 1.57 | 0.000 | 22700 | 3.44 | 1.57 | 2344 | 3.51 | 1.58 | 0.247 |
| **Panel C: Social Connectedness** |  |  |  |  |  |  |  |  |  |  |  |  |  |  |  |
| Feeling Safe | [0] Totally dissatisfied<br>--[10] Totally Satisfied | 27427 | 8.18 | 1.55 | 3015 | 8.31 | 1.56 | 0.002 | 26140 | 8.09 | 1.61 | 2603 | 8.22 | 1.51 | 0.005 |
| Feeling Lonely | [1]-Strongly Disagree<br>- [7] Strongly Agree | 24247 | 2.64 | 1.77 | 2760 | 2.73 | 1.77 | 0.057 | 22603 | 2.74 | 1.74 | 2326 | 2.72 | 1.7 | 0.576 |
| Feeling Part of Local Community | [0] Totally dissatisfied<br>--[10] Totally Satisfied | 27386 | 6.7 | 2.11 | 3012 | 6.65 | 2.2 | 0.275 | 26094 | 6.69 | 2.05 | 2602 | 7 | 1.98 | 0.000 |
| **Panel D: Labor Market and Household Finance** |  |  |  |  |  |  |  |  |  |  |  |  |  |  |  |
| Work Hours (per week in all jobs) | 0-168 | 27445 | 23.93 | 20.18 | 3017 | 20.27 | 20.07 | 0.000 | 26170 | 23.71 | 21.07 | 2604 | 23.78 | 20.37 | 0.912 |
| Weekly Income (current wages & salary in all jobs in \$) | 0-16561 | 27445 | 707.6 | 874.48 | 3017 | 725.51 | 927.45 | 0.392 | 26170 | 717.76 | 909.74 | 2604 | 788.45 | 916.2 | 0.005 |
| Weekly Government Transfer (current transfers excl. family benefits, in \$) | 0-5211 | 27445 | 56.65 | 125.39 | 3017 | 84.48 | 174.33 | 0.000 | 26170 | 61.57 | 132.81 | 2604 | 84.14 | 190.12 | 0.008 |
| **Panel E: Demographics** |  |  |  |  |  |  |  |  |  |  |  |  |  |  |  |
| Age | 15-98 |  | 43.51 | 18.06 |  | 44.04 | 18.39 | 0.215 |  | 42.9 | 18.32 |  | 44.07 | 18.38 | 0.025 |
| Partnered | 0-1 |  | 0.6 | 0.49 |  | 0.61 | 0.49 | 0.410 |  | 0.6 | 0.49 |  | 0.59 | 0.49 | 0.971 |

|  |  |  |  |  |  |  |  |  |  |  |  |
| --- | --- | --- | --- | --- | --- | --- | --- | --- | --- | --- | --- |
| Number of Adults | 0-9 | 2.65 | 1.17 | 2.66 | 1.12 | 0.805 | 2.78 | 1.29 | 2.74 | 1.29 | 0.420 |
| Number of Kids | 0-8 | 0.78 | 1.06 | 0.76 | 1.04 | 0.426 | 0.75 | 1.08 | 0.76 | 1.06 | 0.782 |
| % Graduated from University | 0-1 | 0.33 | 0.47 | 0.35 | 0.48 | 0.094 | 0.37 | 0.48 | 0.41 | 0.49 | 0.060 |

A simple before and during lockdown comparison of outcomes for Melbournians could disguise the possibility that outcome changes were caused by Australia’s first and nation-wide lockdown, which would have also affected individuals in other jurisdictions, or other unobservable factors. For this reason, we opted to compare outcomes for individuals before and during Melbourne’s lockdown period with individuals who resided in Sydney, the most comparable city to Melbourne, where residents were able to move freely. Table 1 shows that Sydneysiders also experienced a decline in mental health, from 72.5 to 70.6 points (*P*<0.001), and in perceptions of feeling part of a community, from 6.7 to 7.0 (*P*<0.001), while their BMIs increased from 25.7 to 26.1 (*P*=0.008). Although Sydneysiders did not change the frequency of physical activity as did Melbournians (*P*=0.247), their perceptions of safety increased from 8.1 to 8.2 (*P*=0.005). In stark contrast to Melbournians, Sydneysiders did not change their workhours (*P*=0.912), but they increased their incomes by 9% from $717.8 per week to $788.5 (*P*=0.005). Thus, the small income changes with workhours remaining constant were most likely due to an increase in the receipt of weekly government transfers by 37% from $61.6 to $84.1 (*P*=0.008).

These findings raised three questions: First, did lockdown cause the changes in the above identified dimensions of human life in Melbourne, or were these driven by other factors that also impacted human life in Sydney? Second, were the changes observed for Melbourne significantly larger in magnitude than the changes in Sydney? Third, if they were larger, were the differences large enough in magnitude to be socially and clinically relevant?

### b. Causal Impact of Lockdown

To answer these questions, we estimated a *difference-in-differences* model that compared outcomes in Melbourne before and during the lockdown with outcomes in Sydney during the same time period (see Methods and Materials). In this econometric model, we controlled for city-level differences and city-specific non-linear time trends in outcomes (since 2011). We furthermore controlled for factors that may have changed over time for individuals: age group, marital status, highest level of education, number of adults in households, number of children in household aged less than 25, and mode and month of interview. Controlling for the interview timing is potentially important, as lockdown may have shifted the interview timing of those residing in Melbourne in 2020. Although we found that the interview schedule distribution hardly changed from previous years, there was a small difference noteworthy for the 2020 interviews: The interviews in Melbourne were completed slightly earlier than in Sydney (fig. S2), while in 2019 rollout dates were identical. This likely reflects Melbournians, locked inside their homes, being easier to reach.

Given the longitudinal nature of the data, we were also able to control for unobserved individual factors that were permanent to the survey participant, but that may have affected outcomes or the way participants reported them. To make the estimates representative for the Australian population, survey-component specific sample weights were applied in each regression. Standard errors were clustered at the household level to adjust for repeated observations within households. All estimates were expressed in terms of standard deviations (*sd*) away from the standardized zero mean (*beta coefficients*).

The model yields a causal estimate of lockdown under the assumption that outcomes in Melbourne and Sydney would have had the same trend in the absence of lockdown. It is common in the literature to test this assumption within a so-called event-study framework, which presents estimated treatment effects before and after policy implementation. We demonstrated that there were no anticipation, or phase-in, effects immediately before the policy was implemented (2018–2019), although for some outcomes trends differed over the full ten years of data (fig. S5). It is for this reason that we controlled flexibly for city-specific time trends (see Materials and Methods).

As much of the discourse has focused upon the inequality of the burden of lockdown, we examined the heterogeneity in the treatment effect by gender, policy-relevant subgroups, (stratified by gender), and exposure length to lockdown. The policy relevant sub-groups were (table S2): (i) whether the household had young children (a proxy for time pressure on and overwork for carers); (ii) whether the respondent lived alone (a proxy for lack of social interaction and loneliness); (iii) whether the respondent had a mental health problem in previous year (a proxy for health care needs and frailty); (iv) whether the household ranked in the bottom household income quintile (a proxy for poverty); and (v) whether the respondent lived in an apartment or semi-detached house (a proxy for lack of over-crowding). To estimate the potential cumulative effects of lockdown, we stratified analyses pragmatically by exposure to lockdown length at the time of the interview: <40 days, 40-70 days, 71-112 days, and days after lockdown was lifted (>112 days). In supplementary material, we present results by age groups.

This econometric model revealed that on average, lockdown had few significant impacts on any domain of human life (fig. 2, tables S3-S4). Regarding the health domain, lockdown led to a significant, but small decline in mental health for women (-0.105sd, *P*=0.043) and a similar, but only marginally significant, decline for men (-0.09sd, *P*=0.087). No significant effects were found for general health or bodily pain.

**Figure 2.**
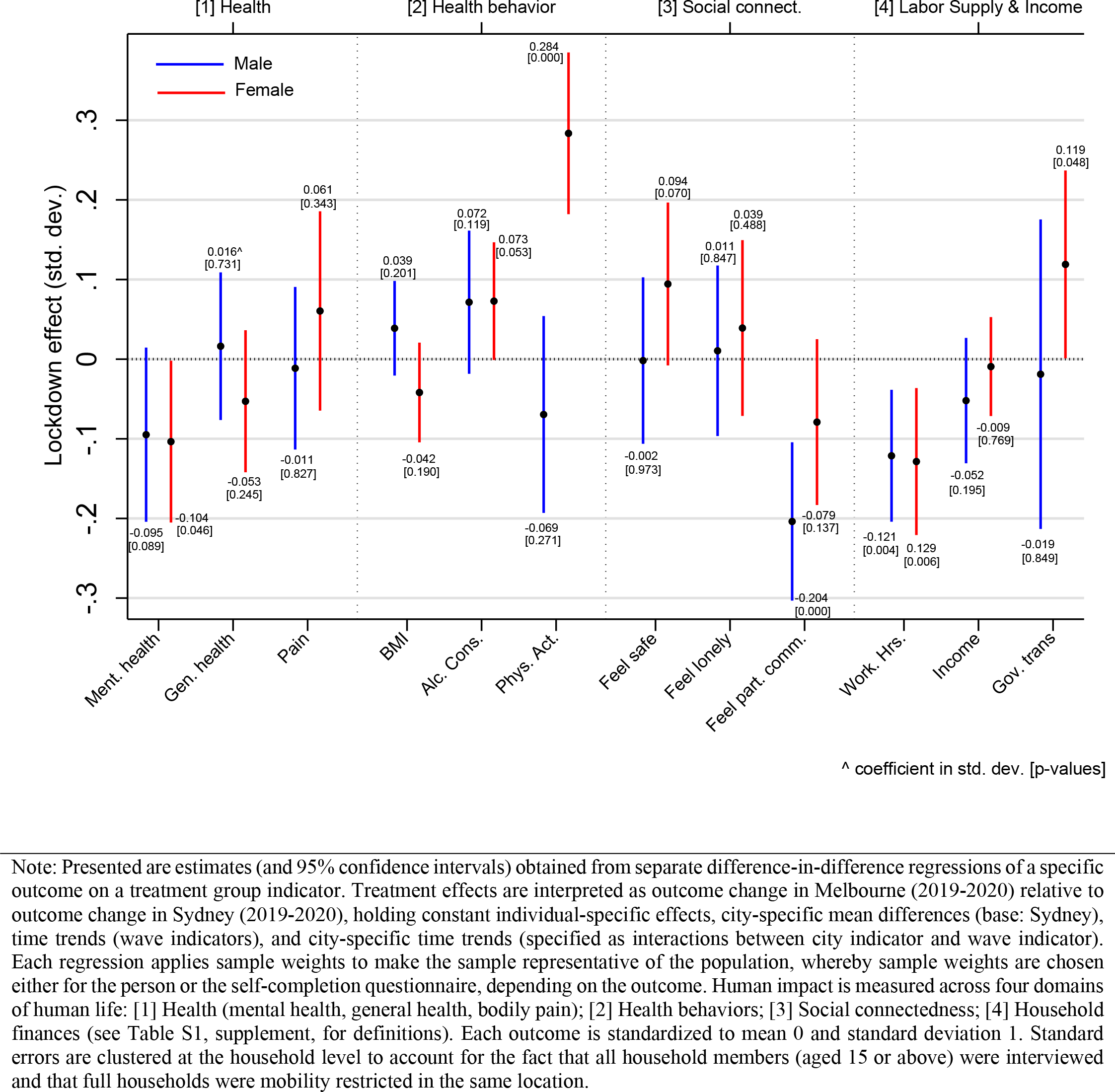
Lockdown treatment effects for 4 domains of human life (expressed in std. dev.), by sex

Regarding health behaviours, lockdown had no significant effects on BMI for either women (BMI: -0.041sd, *P*=0.201) or men (BMI: -0.038sd, *P*=0.206), but we found tentative evidence that lockdown increased the frequency of alcohol consumption of both men (0.071sd, *P*=0.120) and women (0.073sd, *P*=0.054), although the estimates had a large degree of uncertainty and were negligibly small in magnitude (<0.1sd ). Most notable was that lockdown significantly increased the frequency of physical activity for women (0.285sd, *P*<0.001).

In the social connectedness domain we found only one effect: lockdown significantly reduced feelings of being part of the local community for men (-0.204sd, *P*<0.001). Lockdown did not affect current salaries and wages or the amount of government transfers received, but it significantly reduced workhours by -0.121sd (*P*=0.004) for men and -0.129sd (*P*=0.006) for women.

To ensure that our results were not a statistical artefact, we conducted a series of robustness checks that are recommended for differences-in-differences models (see Materials and Methods). The treatment effects were not produced by the 267 survey respondent who were interviewed outside the lockdown period (fig. S6), nor by the choice of the control group (fig. S7). It was also reassuring that we obtained no significant treatment effects when conducting a placebo test, in which we used Sydney as treatment group and the cities of Adelaide, Brisbane and Perth as control group (fig. S8).

### c. Inequalities in the Lockdown Effect

Lockdown had few significant impacts on the average population even when stratified by gender, and even when significant they were mostly small in magnitude (<0.2sd). It is possible, however, that average effects by gender disguised harm to specific groups of the population. We thus re-estimated our model on each outcome by subgroups that we deemed to be most vulnerable to lockdown effects. This heterogeneity analysis revealed that the most vulnerable groups were mothers – women with children younger than age 15 (Fig. 3). Mothers experienced statistically significant and medium-sized declines in mental health (-0.267sd, *P*= 0.014) and general health (-0.228sd, *P*=0.029), and medium-sized increases in feelings of loneliness (0.277sd, *P*=0.020). These negative effects occurred despite increases in feelings of safety (0.264sd, *P*=0.008) and the frequency of physical activity (0.282sd, *P*=0.016). Their workhours decreased by 0.223sd (*P*=0.022), without affecting current salaries (0.000sd, *P*= 0.998) or government transfers (0.035sd, *P*=0.787).

**Figure 3.**
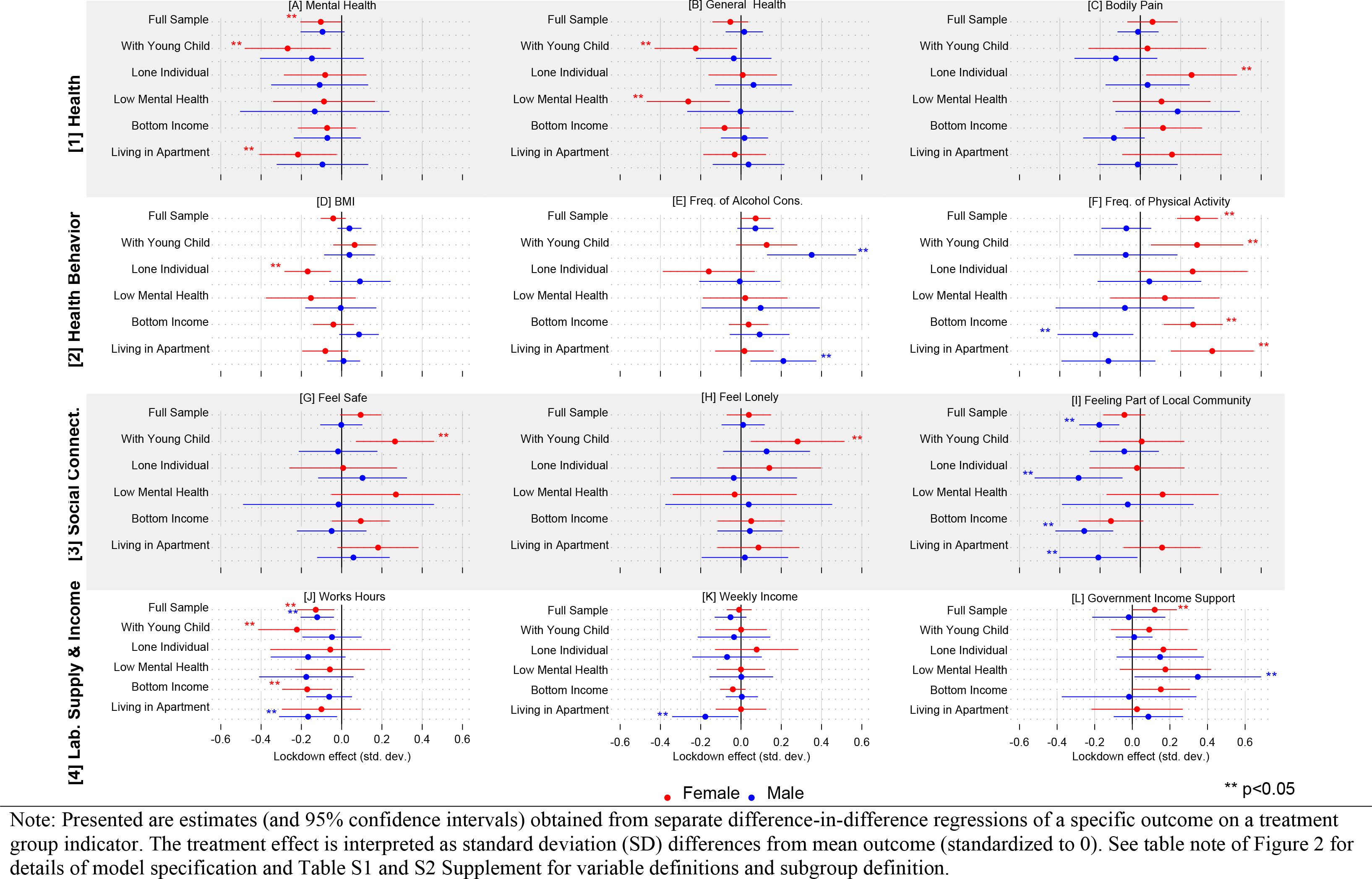
Lockdown treatment effects (expr. in std. dev.), by sex and socially relevant sub-groups

In contrast to mothers, fathers did not experience the same significant lockdown impacts. The only significant impact among fathers was found for the frequency of alcohol consumption, which increased by 0.350sd (*P*=0.002), a medium-sized effect. Fathers also experienced a small drop in mental health (-0.18sd) and an increase in feelings of loneliness (0.16sd), but such effects were not statistically significant. Formal testing of equality of treatment effects between mothers and fathers showed that treatment effect sizes differed significantly in magnitude only for physical activity (*P*=0.032), feelings of safety (*P*=0.032), and marginally for alcohol consumption (*P*=0.075) (table S5). This suggests that fathers may have experienced ill mental health and feelings of loneliness, similar to mothers, although we have uncertainty about the significance of such effects.

We found significant lockdown penalties for men who lived in mobility-restricted dwellings during lockdown. These men felt less socially connected, drank more frequently alcohol, and were harmed in both their hours worked and weekly incomes. In figure 3 we reported lockdown treatment effects by vulnerable groups. We showed that lockdown predominantly harmed mothers with young children. Fathers were also affected to some degree, but not in a statistically significant way.

This does not mean that men were entirely unaffected by lockdown, but they may have been affected in different ways than women with children. The only systematic impact among men was found for men who lived during lockdown in mobility-restricted or smaller area dwellings. Men who lived in apartments or semi-detached houses experienced a significant reduction in workhours (-0.166sd, P=0.024), weekly income (-0.171sd, P=0.040), and perceptions of being part of their community (-0.210sd, P=0.034). This group also increased with moderate effect size the frequency of alcohol consumption (0.206sd) although the effect was not statistically significant (P=0.130).

The question arises whether mobility-restricted living space caused such lockdown penalties, or whether men who live in such dwellings have other characteristics that may have made them more vulnerable economically, socially and behaviorally. As we found no systematic penalties for other men in vulnerable groups - eg the poor, the ones who live alone, or those with previous mental health problems – we can say that it was not poverty, loneliness or health issues that explained the penalties for men in mobility-restricted dwellings. Another possibility is that men who lived in mobility-restricted dwellings were relatively young and healthy, but lockdown harmed them because their lives were most interrupted socially and economically because of less experience in the labor market. Complementary analyses by age groups showed that declines in work hours were indeed driven by younger men (age<35). However, income declines were driven by older men (age 55 or above). It was also both younger and older men who experienced significant declines in feeling part of the local community (figure S9). Hence, there seemed to be heterogeneity among men who live in mobility-restricted dwellings.

Supplementary age group analyses revealed that, with the exception of alcohol use frequency which was driven by middle-aged men (fig. S9E), such effects occurred in the younger age groups (figs. S9I and S9J). We found no ill-health effects of lockdown for the younger or older age groups (figs. S9A and S9B).

### d. Cumulative Lockdown Effects

We also examined whether the length of exposure to lockdown exacerbated the lockdown penalties or benefits. This dose-response analysis revealed that longer exposure to lockdown had no consistent effect across the outcomes; instead, we found heterogeneous trends over exposure time. Fig. 4 (women) and Fig. 5 (men) present our findings, stratified by the presence of young children. For presentation, treatment effects are reported only for selected outcomes and expressed in their original scale to highlight the magnitude of the effects. We tested for equality of estimates between groups at the beginning (<40 days) and the end of the lockdown (71-112 days) (table S6). Because of smaller sample sizes (e.g., the number of observations for mothers in Melbourne during lockdown were: <40 days: 62*n*, 40-70 days: 169*n*, 71-112 days: 53*n*, outside of lockdown: 12*n*), our estimates had large confidence intervals, which affected the reliability of the statistical inference.

**Figure 4.**
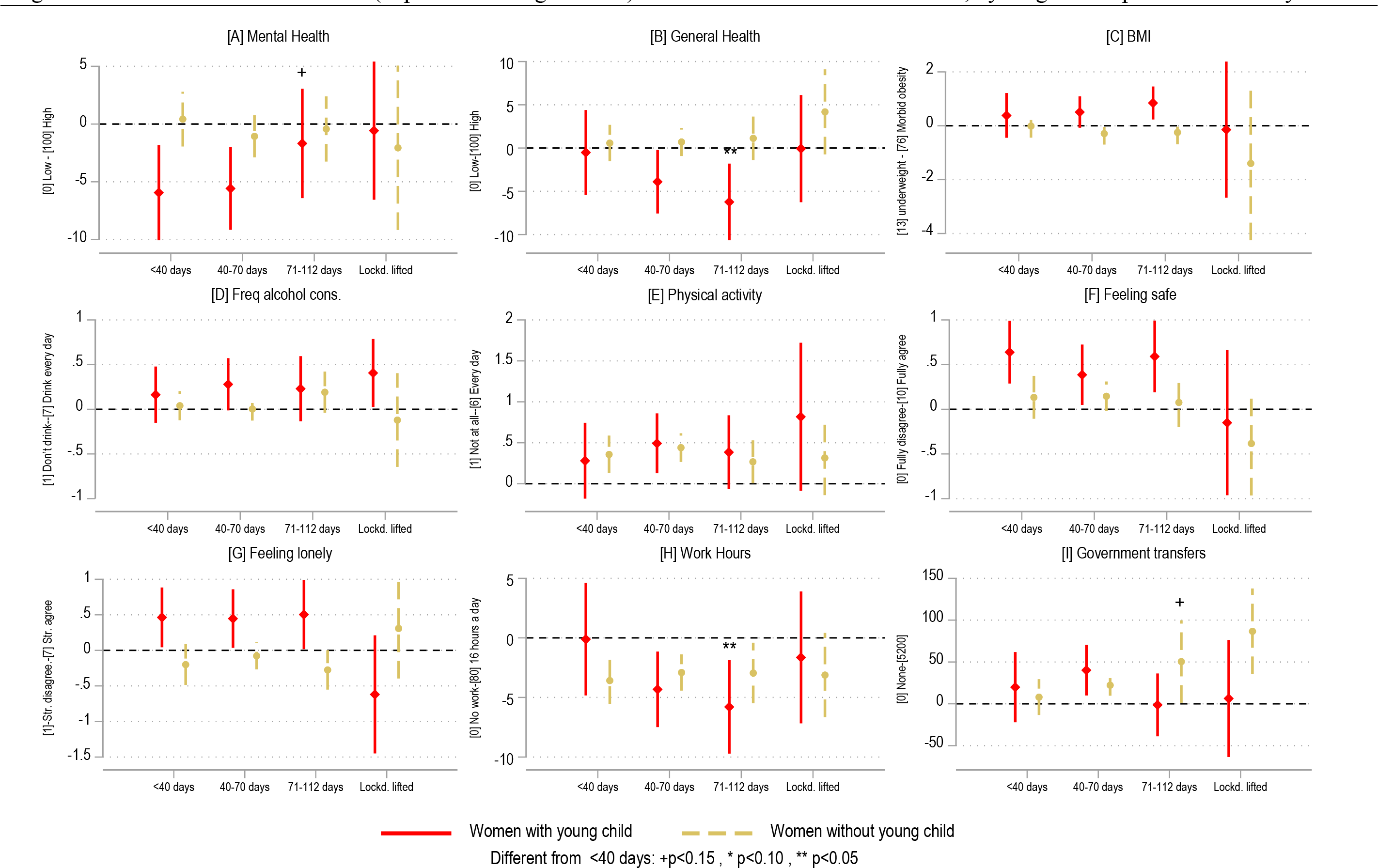
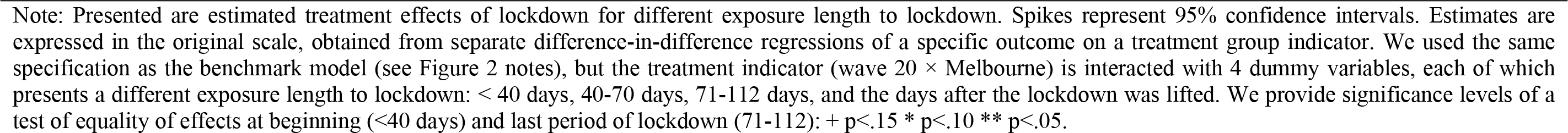
Lockdown treatment effect (expressed in original unit) for women for selected outcomes, by length of exposure and family status

**Figure 5.**
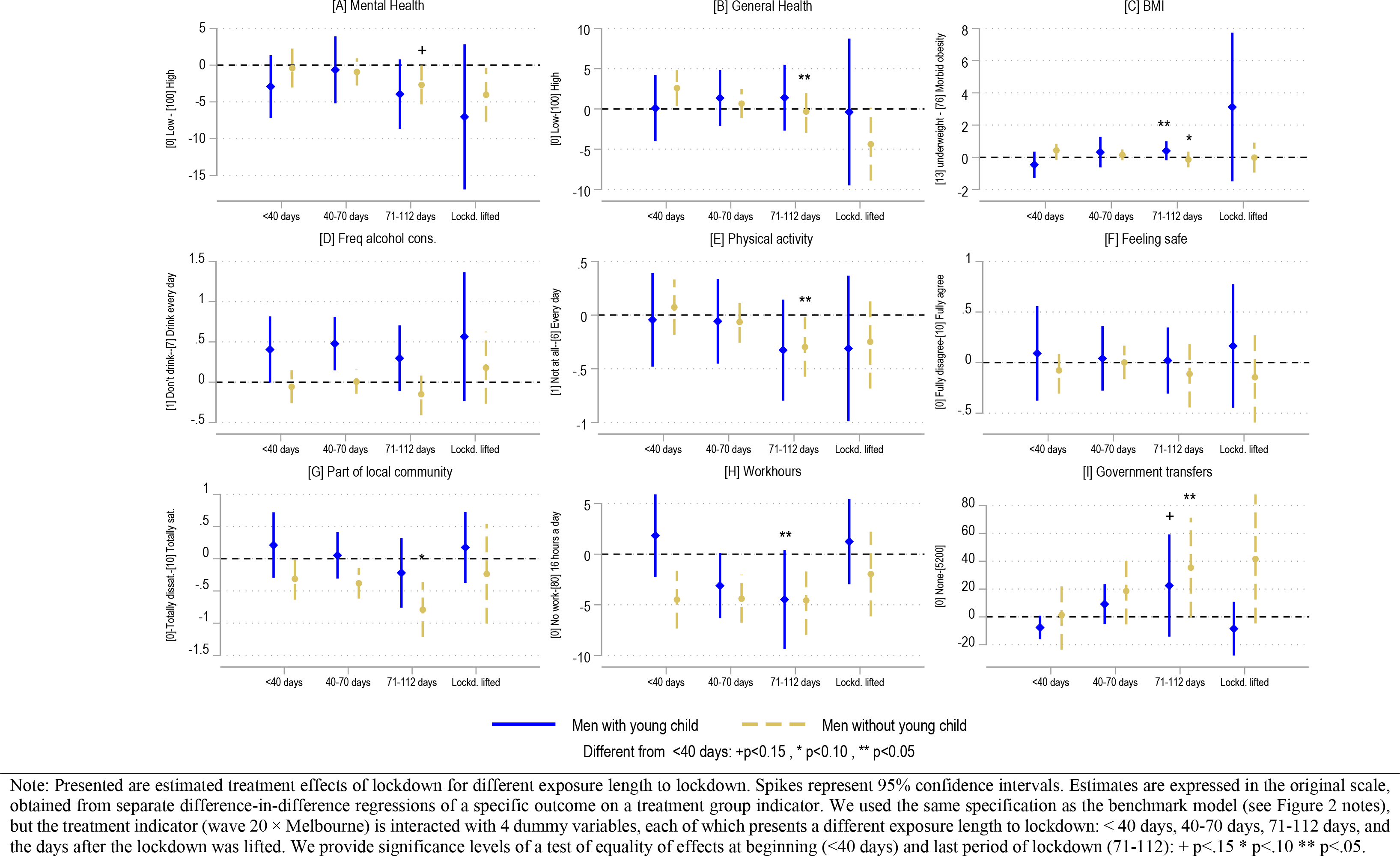
Lockdown treatment effect (expressed in original unit) for men for selected outcomes, by length of exposure and family status

Surprisingly, the mental health penalties of lockdown for mothers faded over time: After plummeting by 6 points (-0.34sd) relative to 2019 values in the early days of lockdown, their mental health scores had returned to their 2019 levels by the end of the lockdown (Fig. 4A, *P*=0.108). In contrast, their general health worsened over the lockdown period. At the beginning of lockdown, we found no difference in general health relative to 2019 levels, but by the end of the lockdown general health was 6 points below 2019 levels (Fig. 4B, *P*=0.030). BMIs tended to increase for mothers, with insignificant effects at the beginning of lockdown but a 1 BMI point increase at the end of lockdown that was significantly different from the 2019 level. However, the difference in BMIs between beginning and the end of lockdown was not statistically significant (*P*=0.282). Workhours per week significantly declined over time as well, with zero treatment effects at the beginning and a 5-hour drop by the end (Fig. 4H, *P*=0.023). Other relevant outcomes were affected uniformly throughout lockdown (alcohol, physical activity, feeling safe).

For fathers, longer lockdown exposure had few additional effects. Their BMIs increased from beginning to the end of lockdown (*P*=0.045). As seen in mothers, fathers reduced their workhours, relative to 2019 levels, only at later stages of the lockdown. Their workhours at the beginning of lockdown were no different to 2019 levels, but fathers worked 5 hours less per week relative to the 2019 levels by the end of lockdown (*P*=0.033). Unlike mothers, the amount of government transfers increased from a zero difference to a difference of A$40 per week relative to 2019 levels, while differences of this treatment effect between beginning and end of lockdown were at the margin of significance (*P*=0.108).

We found more cumulative effects of lockdown for men without children. For them, general health (*P*=0.049), physical activity (*P*=0.027), and feelings of being part of a local community (*P*=0.058) progressively declined.

### e. Could Other Factors Have Explained Our Findings?

Our findings can only be interpreted as causal within the assumptions of the difference-and- difference model. One could argue that the common trend assumption may have been violated if we believed that the onset of the pandemic in Melbourne – roughly 100+ infections per day – could have caused anxiety about the prospects of getting seriously ill, sadness about the actual loss of loved ones, and direct health problems from COVID-19 infection. In supplementary analyses (table S7) we show that such concerns were largely unfounded. While Melbournian men and women were more likely than Sydneysiders to think that they would get infected by (P<0.001) and 9.0% (P<0.001), respectively, which is consistent with expectations of an outbreak with community transmission, they did not have a higher probability of an actual COVID-19 infection at the time of the interview, nor did they say they were more likely to get seriously ill if infected.

We also ruled out that Melbournians entered the second lockdown period significantly more harmed financially by the first lockdown that affected all Australian residents. A difference-in-difference model, which used the same specification as our benchmark model, but used previous financial year incomes (which included incomes up until June 2020) as the outcome variable, showed that Melbournians did not experience greater declines in income relative to the previous financial year than Sydneysiders (table S7). We thus conclude that the quasi-experimental variation exploited in this study was unlikely to have been confounded by differences in pre-lockdown experiences.

## 5. Conclusion

We concluded from our analysis that Zero-COVID policies are not as harmful as may have been expected, but came at high cost to mothers in society, at least in the short run. Women with young children were the group that bore the brunt of the extended lockdown, with estimated penalties that were not small (>0.2sd). Why were mothers so much more strongly affected than other groups? One argument is that mothers carried a greater mental load than usual during lockdown. Mothers are known to work more hours in the household, even in the absence of lockdown. They are more responsible for scheduling, planning and organising family activities (*39*). This cognitive load is likely to become a mental load when this daily process is combined with stress and worry. Mothers during lockdown were more likely to take responsibility for home schooling, which would have added additional workload (31–33). Mothers may also have been affected by the regular presence of their partners. We showed that fathers increased the frequency at which they drank alcohol during lockdown, a risky health behavior. This combined presence of drinking partners in combination with social isolation may have increased family discord, or exposed women to more violence in the home. Concerns over increased interpersonal violence were reported early in context of the costs of lockdown (*41*) and heightened case numbers were expected for Australia by July 2020 (*42*). However, our analysis confirmed that mothers unequivocally felt safer during lockdown. In supplementary analyses we were able to show that mothers and fathers were not significantly less satisfied with their partners (fig. S10). Thus, we ruled out that the harm experienced by mothers was driven by greater risks and partner dissatisfaction in the home. More likely, mothers were simply burdened by additional work hours in the home, which is consistent with a non-compensated, potentially voluntary, reduction in work hours which we observed in our data.

Can our findings be generalized? Our findings were representative of one city in the world which returned to zero new COVID-19 infections through a hard and, at the time, uniquely long lockdown, and which benefitted from relatively strong population compliance. It may be therefore hard to generalize our findings to other jurisdictions and lockdown settings. However, our study design enabled us to rule out the many confounding factors, and we have shown that for mothers the lockdown penalties were mainly present throughout lockdown. We would expect similar results in other countries where lockdown is accompanied by school closures, as was the case in Melbourne. Given the minimal effects observed in other reportedly vulnerable groups this suggests that policy and regulatory responses within lockdowns need to focus on mitigating the effects on mothers of school-aged children.

## Contributors

All authors contributed equally to this paper.

## Declaration of interests

We declare no competing interests.

## Data Availability

All data produced are available online at: https://melbourneinstitute.unimelb.edu.au/hilda

## Acknowledgements

This paper uses unit record data from the Household, Income and Labour Dynamics in Australia (HILDA) Survey, conducted by the Melbourne Institute of Applied Economic and Social Research on behalf of the Australian Government Department of Social Services (DSS) (Release 20, doi:10.26193/PI5LPJ, ADA Dataverse.) The findings and views reported in this paper, however, are those of the authors and should not be attributed to the Australian Government, the DSS, or the Melbourne Institute. The data used are available free of charge to researchers through the National Centre for Longitudinal Data Dataverse at the Australian Data Archive (https://dataverse.ada.edu.au/dataverse/ncld). Access is subject to approval by the Australian Government Department of Social Services and is conditional on signing a license specifying terms of use.

## Supplement: Materials and Methods

### Lockdown rollout

Information on Australia’s pandemic strategy and rollout dates of lockdowns is available in a report of the Australian Institute of Health & Welfare (*1*) (Table 1.2) and a report by the Fair Works Commission (*2*). The JobKeeper program, a fiscal stimulus package that was rolled out early during the first lockdown in Australia, is available in (*3*). Melbourne’s lockdown was facilitated by both voluntary and mandatory mobility restrictions, according to Facebook Data for Good to assess mobility trends in Victoria (*4*). On 1 July, lockdowns were announced for specific postcodes, decreasing mobility substantially in Melbourne (−29%). Stage 4 restrictions were introduced in Greater Melbourne on 1 August. Mobility decreased substantially by −52%. Mobility was significantly less restricted outside the Greater Melbourne area, and it is for this reason we focus the analysis on Greater metropolitan areas (-15% on 1 July 2021 and -34% on 1 Aug 2020, see (*4*)).

### Data

We sourced data from the HILDA Survey. This household panel commenced in 2001 with a nationally representative sample of Australian households (*5*) The first-wave sample comprised 13,969 participants from 7,682 households, which was then followed on an annual basis with all members of those initial households aged 15 years or older, or persons who joined the household later. Individuals gave oral informed consent for participation in the study. Additionally, consent was sought from parents or guardians before seeking the involvement of household members aged less than 18 years. Ethics approval was granted by the Human Research Ethics Committee of the University of Melbourne in 2001 and has been updated or renewed on annual basis since (ID no. 1955879).

In 2011, the sample was refreshed with an additional 2,153 households. Sample loss and attrition were low, with re-interview rates rising from 87% in wave 2 to over 95% by wave 8 and remaining above that level in subsequent waves (*6*).

The data collection process begins every year in August and ends in the following February. Prior to wave 20, more than 90% of interviews were conducted face-to-face. During the pandemic year, 96% of all person interviews in wave 20 were conducted over the phone.

In addition to an individual interview, respondents were asked to complete a separate self-completion questionnaire (SCQ). Prior to wave 20, this was a paper form that was either collected by the interviewer or returned in the mail. In wave 20, an online option was provided, without affecting return rates (in 2020, 91.9%; in 2019, 92.1%).

### Sample

We use 10 waves of data spanning the period 2011 to 2020. After restricting this sample to individuals living in either metropolitan Melbourne or Sydney, the maximum sample size comprised 9,441 individuals (*n*) or 60,108 person-year observations (*pyo*) (Table 1, Stage 1). Analytical sample sizes are slightly smaller due to missing data (60,031 pyo, Stage 2), and vary with the outcome of interest (Stage 3). The final estimation sample included all individuals with at least two repeat observations, ranging between 45,052 *pyo* (alcohol consumption) and 59,236 *pyo* (labor market outcomes) (Stage 4).

### Additional Information on Outcome Measures (see table S1 for all definitions)

We derived from the Medical Outcomes Study Short Form (SF-36 inventory) three measure of health, one of the most widely used self-completion measures of health. These are Mental Health, General Health and Bodily Pain. Previous research has shown that these items in HILDA have good psychometric properties (*7*). These measures were collected in HILDA in the Sefl-Completion Questionnaire, starting in Wave 1.

### Estimation Model

We used methods from the policy evaluation literature (*8–10*) to estimate the causal impact of lockdown on human life outcomes. We exploited the fact that lockdown was implemented only in Melbourne/Victoria and nowhere else in the country, and that individuals could not leave Melbourne/Victoria easily to avoid lockdown rules. The treatment group comprises of individuals who were located in the greater metropolitan area of Melbourne during lockdown. The control group comprised of individuals who were located in the greater metropolitan area of Sydney during the time when Melbourne was locked down. We chose city areas as focus of the analysis, because lockdowns were more likely to be enforced strictly in those areas.

Our models are based on a difference-in-difference specification, which compares changes over time in the treatment group with changes over time in the control group, while holding time trends in outcomes constant in both treatment and control group. We regress outcome Yit on control variables and the treatment indicator:

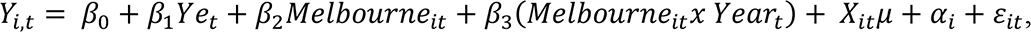

where *Melbourne*_*it*_ equals 1 if the respondent lived in Melbourne in year “t” and 0 in Sydney. *Year*_*t*_ includes year fixed effects leaving 2019 as the base category. Our coefficient of interest is included in β_3_which reports the interaction between our “treatment” variable (i.e., being Melbourne) and the wave (i.e., time) dummies. For example, if the longer lockdown has any impact on mental health such interaction should come out significant at year 2020. Additionally, in order to comply with the parallel trends assumption, the interaction coefficient should not be significant in the waves prior to the pandemic. Otherwise, this would indicate that mental health trends had already followed different trajectories across cities prior to the divergence in containment policies. *X*_*it*_ is a vector of control variables including age as categorical variable, number of individuals, number of children, education, marital status (all dummy variables). We also control, the month and mode of interview.

As an alternative specification, we estimate a fixed effect model in which we defined treatment group as people live in Melbourne in 2020 and control group as people live in Sydney in 2020. Formally, we estimate

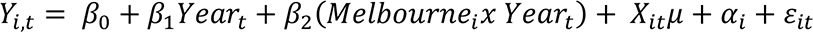

where *Melbourne*_*i*_ equals 1 if the respondent lived in Melbourne in 2020 (when HILDA collected wave 20 information) and 0 in Sydney. Note that in this model, it is not necessary to include the treatment dummy (*Melbourne*_*it*_) alone since it is fixed over time and therefore will be absorbed by the individual fixed effects. *Year*_*t*_ includes year fixed effects leaving 2019 as the base category. In this model the parameter of interest is β_2_. Our results from this model are similar to the above model presented in the paper.

All models use survey-component specific sample probability weights to make the samples representative for the population in each wave. Standard errors are clustered at the household level, to adjust for non-independent observations per household.

All estimates are reported as standardized coefficients, which is interpreted as a percent standard deviation difference in outcomes before and during lockdown for the treatment group relative to the control group.

As is common in the difference-in-difference literature, one needs to test the common trend assumptions of the model. This can be done by presenting so-called event-study graphs, alternative parametrization of trends for treatment and control groups, and so-called placebo tests (*10*).

## Supplement: Tables and Figures

**Table S1.** Definition, scaling, and transformation of outcome variables

| Outcome | Question/Definition | Original Categories | Transformed Categories |
| --- | --- | --- | --- |
| *Panel A: Health* |  |  |  |
| [1] Mental Health | SF-36 mental health - transformed | 0-100 |  |
| [2] General Health | SF-36 general health - transformed | 0-100 |  |
| [3] Bodily Pain | SF-36 bodily pain - transformed | 0-100 |  |
| *Panel B: Health behaviors* |  |  |  |
| [4] Body Mass Index | Weight in kilograms divided by square of height in meters |  |  |
| [5] Frequency of Alcohol Consumption | Frequency of Alcohol Drinking | [1] I have never drunk alcohol<br>[2] I no longer drink<br>[3] Yes, I drink alcohol everyday<br>[4] Yes, I drink alcohol 5 or 6 days per week<br>[5] Yes, I drink alcohol 3 or 4 days per week<br>[6] Yes, I drink alcohol 1 or 2 days per week<br>[7] Yes, I drink alcohol 2 or 3 days per month<br>[8] Yes, but only rarely | Exclude people who state “have never drunk alcohol”<br>[1] I no longer drink<br>[2] Yes, but only rarely<br>[3] Yes, I drink alcohol 2 or 3 days per month<br>[4] Yes, I drink alcohol 1 or 2 days per week<br>[5] Yes, I drink alcohol 3 or 4 days per week<br>[6] Yes, I drink alcohol 5 or 6 days per week<br>[7] Yes, I drink alcohol everyday |
| [6] Frequency of Physical Activity | In general, how often do you participate in moderate or intensive physical activity for at least 30 minutes | [1] Not at all<br>[2] Less than once a week<br>[3] 1 to 2 times a week<br>[4] 3 times a week<br>[5] More than 3 times a week<br>[6] Every day | [1] Not at all<br>[2] Less than once a week<br>[3] 1 to 2 times a week<br>[4] 3 times a week<br>[5] More than 3 times a week<br>[6] Every day |

| Outcome | Definition | Original Categories | Transformed Categories |
| --- | --- | --- | --- |
| *Panel C: Social Connectedness* |  |  |  |
| [7] Feeling Safe | How Safe you feel | (0-Totally dissatisfied, 10-Totally Satisfied) | (0-Totally dissatisfied, 10- Totally Satisfied) |
| [8] Feeling Lonely | I often feel very lonely | lonely (1-Strongly Disagree; 7- Strongly Agree) | lonely (1-Strongly Disagree; 7- Strongly Agree) |
| [9] Feeling Part of Local Community | Feeling part of your local community | (0-Totally dissatisfied, 10-Totally Satisfied) | (0-Totally dissatisfied, 10- Totally Satisfied) |
| *Panel D: Labor market and household finance* |  |  |  |
| [10] Work Hour | Hours per week usually worked in all jobs | Responding Person – Derived Variable | 0-168 |
| [11] Weekly Income | Current weekly gross wages & salary - all jobs (\$) imputed | Responding Person – Derived Variable | 0-16561 |
| [12] Weekly Government Transfer | Current weekly Australian public transfers excluding family benefits (\$) [imputed] | Responding Person – Derived Variable | 0-5211 |

**Table S2.**
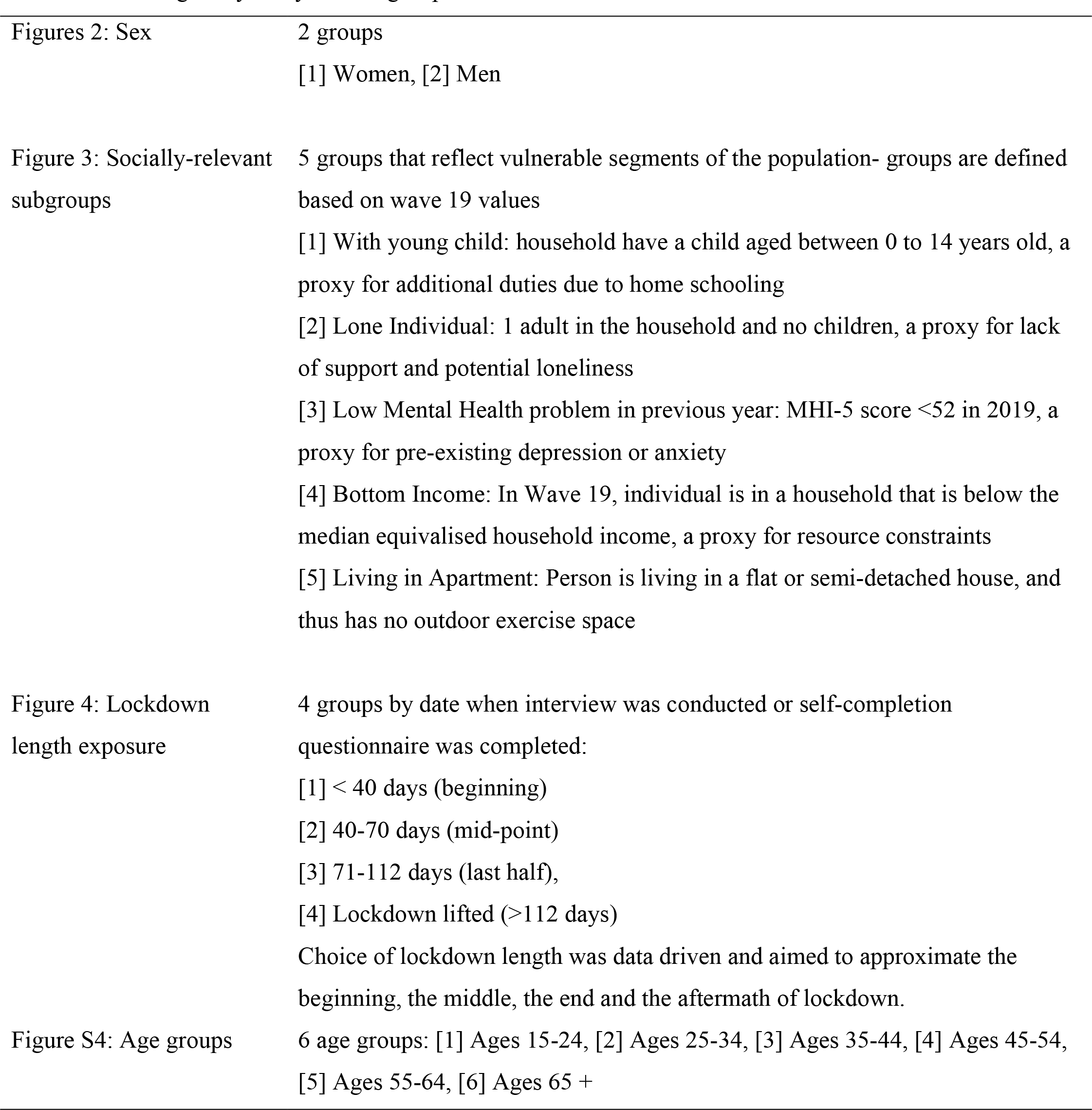
Heterogeneity analysis: sub-group definitions

**Table S3.** Full estimation results for health outcomes and health behavior (estimated treatment effects expressed in standardised to mean 0, std. dev. 1)

|  | Mental Health |  | General Health |  | Bodily Pain |  | BMI |  | Alcohol Freq. |  | Exercise Freq. |  |
| --- | --- | --- | --- | --- | --- | --- | --- | --- | --- | --- | --- | --- |
|  | Male | Female | Male | Female | Male | Female | Male | Female | Male | Female | Male | Female |
|  | (1) | (2) | (3) | (4) | (5) | (6) | (7) | (8) | (9) | (10) | (11) | (12) |
| **City (Sydney is base category)** |  |  |  |  |  |  |  |  |  |  |  |  |
| Melbourne | 0.001<br>(0.115) | 0.230*<br>(0.129) | -0.066<br>(0.111) | 0.102<br>(0.119) | 0.142<br>(0.121) | -0.172<br>(0.109) | -0.004<br>(0.043) | -0.014<br>(0.057) | 0.019<br>(0.105) | -0.052<br>(0.112) | 0.133<br>(0.149) | 0.034<br>(0.116) |
| **Wave** |  |  |  |  |  |  |  |  |  |  |  |  |
| Wave 11 | -0.009<br>(0.055) | 0.052<br>(0.044) | 0.188***<br>(0.043) | 0.227***<br>(0.037) | 0.152***<br>(0.054) | 0.111**<br>(0.052) | -0.119***<br>(0.027) | -0.175***<br>(0.023) | 0.046<br>(0.041) | -0.025<br>(0.032) | 0.102*<br>(0.054) | 0.006<br>(0.047) |
| Wave 12 | 0.103*<br>(0.058) | 0.016<br>(0.048) | 0.187***<br>(0.040) | 0.193***<br>(0.044) | 0.178***<br>(0.050) | 0.110**<br>(0.047) | -0.094***<br>(0.028) | -0.156***<br>(0.023) | 0.001<br>(0.036) | -0.002<br>(0.034) | 0.063<br>(0.058) | 0.033<br>(0.048) |
| Wave 13 | 0.037<br>(0.046) | 0.008<br>(0.047) | 0.169***<br>(0.038) | 0.221***<br>(0.036) | 0.135***<br>(0.046) | 0.133***<br>(0.048) | -0.086***<br>(0.024) | -0.141***<br>(0.023) | 0.010<br>(0.037) | -0.047<br>(0.030) | 0.090*<br>(0.048) | 0.061<br>(0.047) |
| Wave 14 | 0.085*<br>(0.045) | -0.020<br>(0.050) | 0.127***<br>(0.039) | 0.128***<br>(0.035) | 0.162***<br>(0.046) | 0.021<br>(0.057) | -0.079***<br>(0.021) | -0.124***<br>(0.023) | 0.008<br>(0.034) | -0.042<br>(0.030) | 0.014<br>(0.048) | -0.095**<br>(0.043) |
| Wave 15 | 0.045<br>(0.049) | 0.007<br>(0.042) | 0.131***<br>(0.037) | 0.096***<br>(0.035) | 0.117**<br>(0.048) | 0.052<br>(0.044) | -0.066***<br>(0.020) | -0.097***<br>(0.023) | -0.020<br>(0.040) | -0.034<br>(0.029) | 0.006<br>(0.047) | -0.101**<br>(0.046) |
| Wave 16 | 0.040<br>(0.050) | -0.052<br>(0.045) | 0.130***<br>(0.037) | 0.105***<br>(0.036) | 0.075*<br>(0.041) | 0.013<br>(0.043) | -0.053***<br>(0.019) | -0.051***<br>(0.019) | -0.011<br>(0.033) | 0.015<br>(0.031) | 0.021<br>(0.044) | -0.078*<br>(0.043) |
| Wave 17 | 0.008<br>(0.052) | -0.009<br>(0.044) | 0.076**<br>(0.037) | 0.084**<br>(0.037) | 0.063<br>(0.043) | -0.023<br>(0.045) | -0.019<br>(0.021) | -0.034*<br>(0.019) | -0.010<br>(0.033) | -0.012<br>(0.027) | -0.023<br>(0.047) | -0.062<br>(0.038) |
| Wave 18 | -0.008<br>(0.044) | -0.000<br>(0.041) | 0.066*<br>(0.035) | 0.063*<br>(0.038) | 0.016<br>(0.040) | -0.026<br>(0.041) | -0.012<br>(0.019) | -0.034*<br>(0.020) | -0.004<br>(0.036) | 0.020<br>(0.027) | 0.044<br>(0.043) | -0.043<br>(0.043) |
| Wave 19 (base) | 0.000<br>(.) | 0.000<br>(.) | 0.000<br>(.) | 0.000<br>(.) | 0.000<br>(.) | 0.000<br>(.) | 0.000<br>(.) | 0.000<br>(.) | 0.000<br>(.) | 0.000<br>(.) | 0.000<br>(.) | 0.000<br>(.) |
| Wave 20 | -0.007<br>(0.054) | 0.005<br>(0.051) | 0.023<br>(0.053) | 0.099**<br>(0.045) | 0.016<br>(0.057) | -0.030<br>(0.060) | 0.003<br>(0.027) | 0.066**<br>(0.029) | -0.015<br>(0.044) | -0.009<br>(0.040) | -0.043<br>(0.063) | -0.022<br>(0.050) |
| **Melbourne X Wave** |  |  |  |  |  |  |  |  |  |  |  |  |
| Melbourne X Wave 11 | 0.090<br>(0.060) | 0.022<br>(0.055) | -0.001<br>(0.048) | -0.057<br>(0.047) | -0.065<br>(0.061) | 0.185***<br>(0.063) | -0.034<br>(0.031) | -0.020<br>(0.029) | -0.006<br>(0.049) | 0.053<br>(0.038) | -0.064<br>(0.063) | 0.100*<br>(0.056) |
| Melbourne X Wave 12 | -0.059<br>(0.064) | 0.080<br>(0.057) | -0.018<br>(0.046) | -0.014<br>(0.052) | -0.073<br>(0.057) | 0.112**<br>(0.053) | -0.050<br>(0.032) | -0.021<br>(0.027) | 0.016<br>(0.045) | -0.008<br>(0.039) | -0.030<br>(0.067) | 0.058<br>(0.055) |
| Melbourne X Wave 13 | 0.055<br>(0.055) | 0.110**<br>(0.054) | -0.022<br>(0.044) | -0.056<br>(0.045) | -0.080<br>(0.055) | 0.096*<br>(0.055) | -0.019<br>(0.029) | -0.025<br>(0.029) | 0.027<br>(0.046) | 0.023<br>(0.037) | -0.131**<br>(0.059) | 0.038<br>(0.055) |
| Melbourne X Wave 14 | -0.043<br>(0.053) | 0.112*<br>(0.058) | -0.052<br>(0.046) | -0.007<br>(0.043) | -0.091*<br>(0.055) | 0.148**<br>(0.063) | -0.033<br>(0.026) | -0.038<br>(0.028) | 0.011<br>(0.043) | 0.053<br>(0.035) | -0.053<br>(0.060) | 0.141***<br>(0.051) |
| Melbourne X Wave 15 | -0.013<br>(0.057) | 0.078<br>(0.052) | -0.059<br>(0.045) | 0.019<br>(0.044) | -0.076<br>(0.058) | 0.128**<br>(0.052) | -0.027<br>(0.025) | -0.042<br>(0.030) | 0.048<br>(0.051) | 0.046<br>(0.036) | -0.099<br>(0.061) | 0.132**<br>(0.057) |
| Melbourne X Wave 16 | -0.025<br>(0.059) | 0.151***<br>(0.054) | -0.065<br>(0.044) | -0.037<br>(0.044) | -0.012<br>(0.050) | 0.128**<br>(0.052) | -0.001<br>(0.025) | -0.063**<br>(0.025) | 0.062<br>(0.043) | 0.012<br>(0.037) | -0.052<br>(0.058) | 0.109**<br>(0.052) |
| Melbourne X Wave 17 | -0.009<br>(0.059) | 0.060<br>(0.053) | -0.021<br>(0.044) | -0.044<br>(0.045) | -0.046<br>(0.052) | 0.123**<br>(0.056) | -0.009<br>(0.026) | -0.044*<br>(0.025) | 0.042<br>(0.043) | 0.049<br>(0.033) | -0.024<br>(0.061) | 0.103**<br>(0.049) |
| Melbourne X Wave 18 | 0.017<br>(0.052) | 0.030<br>(0.051) | -0.024<br>(0.041) | -0.033<br>(0.045) | -0.013<br>(0.051) | 0.066<br>(0.052) | -0.005<br>(0.026) | -0.015<br>(0.027) | 0.020<br>(0.047) | 0.004<br>(0.034) | -0.102*<br>(0.059) | 0.034<br>(0.052) |
| Melbourne X Wave 19 (base) | 0.000<br>(.) | 0.000<br>(.) | 0.000<br>(.) | 0.000<br>(.) | 0.000<br>(.) | 0.000<br>(.) | 0.000<br>(.) | 0.000<br>(.) | 0.000<br>(.) | 0.000<br>(.) | 0.000<br>(.) | 0.000<br>(.) |
| Melbourne X Wave 20 | -0.095* | -0.104** | 0.016 | -0.053 | -0.011 | 0.061 | 0.039 | -0.042 | 0.072 | 0.073* | -0.069 | 0.284*** |
|  | (0.056) | (0.052) | (0.047) | (0.045) | (0.052) | (0.064) | (0.030) | (0.032) | (0.046) | (0.038) | (0.063) | (0.052) |
| **Age Groups** |  |  |  |  |  |  |  |  |  |  |  |  |
| 14-17 | 0.270*** | 0.225** | 0.181*** | 0.105 | -0.011 | -0.011 | -0.359*** | -0.276*** | -0.524*** | -0.304*** | 0.359*** | 0.056 |
|  | (0.089) | (0.098) | (0.066) | (0.073) | (0.079) | (0.119) | (0.045) | (0.059) | (0.064) | (0.063) | (0.083) | (0.105) |
| 18-24 | 0.062 | 0.029 | 0.022 | -0.065* | -0.009 | -0.125** | -0.087*** | -0.091*** | -0.041 | 0.005 | 0.153*** | -0.135*** |
|  | (0.045) | (0.049) | (0.034) | (0.035) | (0.047) | (0.057) | (0.022) | (0.024) | (0.031) | (0.026) | (0.047) | (0.045) |
| 25-34 (base) | 0.000 | 0.000 | 0.000 | 0.000 | 0.000 | 0.000 | 0.000 | 0.000 | 0.000 | 0.000 | 0.000 | 0.000 |
|  | (.) | (.) | (.) | (.) | (.) | (.) | (.) | (.) | (.) | (.) | (.) | (.) |
| 35-44 | -0.067 | -0.063 | -0.017 | -0.026 | 0.027 | 0.051 | 0.047** | -0.002 | 0.004 | 0.022 | -0.072 | 0.086** |
|  | (0.047) | (0.041) | (0.036) | (0.043) | (0.040) | (0.046) | (0.020) | (0.023) | (0.044) | (0.032) | (0.046) | (0.042) |
| 45-54 | -0.050 | -0.136** | -0.055 | -0.056 | 0.017 | 0.028 | 0.053* | -0.046 | 0.022 | -0.014 | 0.032 | 0.103* |
|  | (0.061) | (0.061) | (0.050) | (0.056) | (0.058) | (0.065) | (0.029) | (0.032) | (0.057) | (0.046) | (0.068) | (0.061) |
| 55-64 | -0.021 | -0.093 | -0.112 | -0.049 | -0.017 | 0.077 | 0.038 | -0.060 | 0.053 | 0.013 | 0.029 | 0.201** |
|  | (0.084) | (0.074) | (0.068) | (0.070) | (0.081) | (0.081) | (0.039) | (0.038) | (0.066) | (0.057) | (0.087) | (0.079) |
| 65-74 | 0.063 | 0.022 | -0.134 | 0.000 | -0.066 | 0.136 | -0.016 | -0.157*** | -0.025 | -0.057 | 0.175 | 0.266*** |
|  | (0.101) | (0.088) | (0.083) | (0.082) | (0.101) | (0.098) | (0.049) | (0.047) | (0.080) | (0.068) | (0.111) | (0.098) |
| 75-84 | 0.039 | 0.033 | -0.213** | -0.095 | -0.176 | 0.110 | -0.148** | -0.304*** | -0.149 | -0.194** | 0.074 | 0.124 |
|  | (0.119) | (0.113) | (0.098) | (0.107) | (0.122) | (0.122) | (0.058) | (0.060) | (0.095) | (0.083) | (0.136) | (0.123) |
| 85+ | -0.241 | -0.059 | -0.653*** | -0.217* | -0.477*** | 0.025 | -0.248*** | -0.452*** | -0.226 | -0.408*** | -0.473** | -0.172 |
|  | (0.148) | (0.135) | (0.139) | (0.124) | (0.166) | (0.151) | (0.076) | (0.074) | (0.142) | (0.114) | (0.209) | (0.155) |
| **Marital Status** |  |  |  |  |  |  |  |  |  |  |  |  |
| Legally Married (base) | 0.000 | 0.000 | 0.000 | 0.000 | 0.000 | 0.000 | 0.000 | 0.000 | 0.000 | 0.000 | 0.000 | 0.000 |
|  | (.) | (.) | (.) | (.) | (.) | (.) | (.) | (.) | (.) | (.) | (.) | (.) |
| In de-facto relationship | 0.053<br>(0.042) | -0.043<br>(0.053) | 0.028<br>(0.037) | 0.005<br>(0.033) | -0.010<br>(0.045) | -0.005<br>(0.065) | 0.008<br>(0.026) | -0.103***<br>(0.024) | -0.001<br>(0.028) | 0.126***<br>(0.030) | -0.014<br>(0.044) | 0.095**<br>(0.048) |
| Separated | -0.220**<br>(0.087) | -0.290***<br>(0.065) | -0.235***<br>(0.078) | -0.024<br>(0.056) | -0.107<br>(0.102) | 0.122<br>(0.090) | -0.104***<br>(0.032) | -0.095***<br>(0.036) | -0.051<br>(0.056) | 0.131**<br>(0.055) | -0.085<br>(0.145) | 0.151*<br>(0.083) |
| Divorced | -0.013<br>(0.106) | -0.089<br>(0.081) | -0.187<br>(0.154) | 0.069<br>(0.053) | -0.305<br>(0.196) | 0.117<br>(0.079) | -0.059<br>(0.039) | -0.135***<br>(0.030) | -0.110**<br>(0.053) | 0.161***<br>(0.050) | 0.224*<br>(0.115) | 0.122*<br>(0.071) |
| Widowed | -0.067<br>(0.102) | -0.086<br>(0.075) | 0.034<br>(0.084) | -0.036<br>(0.058) | -0.064<br>(0.106) | -0.030<br>(0.078) | -0.099*<br>(0.059) | -0.041<br>(0.040) | -0.060<br>(0.103) | -0.007<br>(0.055) | -0.070<br>(0.103) | -0.037<br>(0.085) |
| Single | -0.052<br>(0.059) | -0.097<br>(0.061) | 0.003<br>(0.053) | -0.061<br>(0.043) | -0.010<br>(0.061) | -0.016<br>(0.066) | -0.028<br>(0.040) | -0.242***<br>(0.030) | 0.018<br>(0.041) | 0.228***<br>(0.040) | 0.155**<br>(0.062) | 0.081<br>(0.059) |
| **Highest Education Achieved** |  |  |  |  |  |  |  |  |  |  |  |  |
| Less than Year 12 | -0.020<br>(0.101) | 0.069<br>(0.080) | -0.153**<br>(0.077) | -0.152**<br>(0.073) | -0.118<br>(0.095) | -0.256<br>(0.168) | -0.045<br>(0.046) | -0.024<br>(0.052) | -0.152**<br>(0.072) | -0.102*<br>(0.056) | -0.086<br>(0.082) | -0.038<br>(0.129) |
| Year 12 | 0.032<br>(0.068) | 0.114**<br>(0.051) | -0.032<br>(0.059) | 0.001<br>(0.038) | -0.037<br>(0.075) | 0.003<br>(0.054) | -0.032<br>(0.034) | -0.058**<br>(0.025) | -0.203***<br>(0.052) | -0.073**<br>(0.037) | -0.105*<br>(0.063) | 0.089<br>(0.057) |
| Certificate | -0.034<br>(0.099) | 0.140<br>(0.098) | -0.151*<br>(0.081) | 0.029<br>(0.056) | -0.081<br>(0.099) | 0.026<br>(0.102) | 0.056<br>(0.051) | 0.023<br>(0.035) | -0.190**<br>(0.077) | -0.111**<br>(0.045) | -0.170*<br>(0.092) | 0.043<br>(0.093) |
| Diploma | -0.137<br>(0.156) | 0.058<br>(0.086) | -0.041<br>(0.081) | 0.034<br>(0.063) | -0.053<br>(0.127) | -0.046<br>(0.089) | 0.100<br>(0.076) | 0.151**<br>(0.059) | -0.222**<br>(0.093) | -0.084<br>(0.054) | 0.126<br>(0.165) | 0.058<br>(0.082) |
| Bachelor degree (base) | 0.000<br>(.) | 0.000<br>(.) | 0.000<br>(.) | 0.000<br>(.) | 0.000<br>(.) | 0.000<br>(.) | 0.000<br>(.) | 0.000<br>(.) | 0.000<br>(.) | 0.000<br>(.) | 0.000<br>(.) | 0.000<br>(.) |
| Higher than Bachelor | -0.074<br>(0.082) | -0.032<br>(0.073) | -0.106*<br>(0.064) | 0.016<br>(0.052) | 0.058<br>(0.068) | -0.015<br>(0.099) | 0.024<br>(0.035) | 0.010<br>(0.024) | -0.007<br>(0.049) | 0.036<br>(0.067) | 0.108<br>(0.076) | 0.060<br>(0.068) |
| **Interview Month** |  |  |  |  |  |  |  |  |  |  |  |  |
| August (base) | 0.000 | 0.000 | 0.000 | 0.000 | 0.000 | 0.000 | 0.000 | 0.000 | 0.000 | 0.000 | 0.000 | 0.000 |

|  | (.) | (.) | (.) | (.) | (.) | (.) | (.) | (.) | (.) | (.) | (.) | (.) |
| --- | --- | --- | --- | --- | --- | --- | --- | --- | --- | --- | --- | --- |
| September | 0.098<br>(0.124) | -0.008<br>(0.148) | 0.028<br>(0.098) | -0.047<br>(0.091) | -0.209*<br>(0.112) | 0.054<br>(0.089) | -0.019<br>(0.046) | -0.011<br>(0.063) | -0.077<br>(0.075) | -0.085<br>(0.103) | 0.152<br>(0.136) | -0.162<br>(0.140) |
| October | 0.120<br>(0.111) | -0.077<br>(0.076) | -0.063<br>(0.066) | -0.119*<br>(0.068) | -0.190*<br>(0.105) | 0.021<br>(0.082) | 0.004<br>(0.045) | -0.003<br>(0.053) | -0.039<br>(0.061) | 0.014<br>(0.054) | 0.092<br>(0.093) | -0.298***<br>(0.110) |
| November | 0.159*<br>(0.082) | -0.072<br>(0.058) | -0.008<br>(0.045) | -0.102**<br>(0.049) | -0.110<br>(0.071) | -0.069<br>(0.052) | 0.002<br>(0.032) | 0.054<br>(0.045) | -0.049<br>(0.047) | 0.011<br>(0.041) | 0.064<br>(0.064) | -0.202**<br>(0.080) |
| December | 0.118<br>(0.081) | -0.121**<br>(0.058) | -0.021<br>(0.045) | -0.093*<br>(0.048) | -0.111<br>(0.071) | -0.134***<br>(0.052) | -0.002<br>(0.033) | 0.056<br>(0.045) | -0.034<br>(0.047) | 0.017<br>(0.041) | 0.054<br>(0.064) | -0.200**<br>(0.079) |
| January | 0.138*<br>(0.083) | -0.090<br>(0.060) | -0.033<br>(0.048) | -0.108**<br>(0.051) | -0.115<br>(0.073) | -0.107*<br>(0.058) | 0.011<br>(0.034) | 0.052<br>(0.049) | -0.042<br>(0.049) | -0.009<br>(0.044) | 0.065<br>(0.068) | -0.195**<br>(0.081) |
| February | 0.103<br>(0.087) | -0.137**<br>(0.066) | -0.045<br>(0.055) | -0.145**<br>(0.058) | -0.148*<br>(0.079) | -0.104*<br>(0.060) | 0.044<br>(0.038) | 0.069<br>(0.046) | 0.042<br>(0.059) | 0.042<br>(0.044) | 0.142*<br>(0.077) | -0.176**<br>(0.083) |
| March | 0.194*<br>(0.110) | -0.104<br>(0.105) | 0.095<br>(0.074) | -0.047<br>(0.078) | 0.016<br>(0.113) | -0.157<br>(0.101) | -0.011<br>(0.044) | 0.006<br>(0.060) | -0.014<br>(0.083) | 0.073<br>(0.067) | -0.000<br>(0.113) | -0.284**<br>(0.130) |
| **No of Adults** |  |  |  |  |  |  |  |  |  |  |  |  |
| 1 (base) | 0.000<br>(.) | 0.000<br>(.) | 0.000<br>(.) | 0.000<br>(.) | 0.000<br>(.) | 0.000<br>(.) | 0.000<br>(.) | 0.000<br>(.) | 0.000<br>(.) | 0.000<br>(.) | 0.000<br>(.) | 0.000<br>(.) |
| 2 | 0.063<br>(0.043) | -0.038<br>(0.035) | 0.038<br>(0.040) | -0.041<br>(0.026) | -0.065<br>(0.044) | -0.048<br>(0.035) | -0.060**<br>(0.026) | -0.001<br>(0.018) | -0.000<br>(0.032) | 0.015<br>(0.024) | -0.020<br>(0.047) | -0.001<br>(0.037) |
| 3 | 0.107**<br>(0.046) | -0.040<br>(0.042) | 0.044<br>(0.044) | -0.049<br>(0.030) | -0.055<br>(0.049) | -0.031<br>(0.040) | -0.065**<br>(0.026) | -0.017<br>(0.021) | 0.011<br>(0.036) | 0.018<br>(0.025) | 0.079<br>(0.055) | 0.073*<br>(0.042) |
| 4 | 0.105**<br>(0.047) | 0.007<br>(0.046) | 0.003<br>(0.045) | -0.105***<br>(0.037) | -0.053<br>(0.050) | -0.077<br>(0.047) | -0.029<br>(0.028) | -0.023<br>(0.024) | -0.014<br>(0.039) | -0.009<br>(0.028) | 0.002<br>(0.054) | 0.055<br>(0.045) |
| 5 | 0.068<br>(0.059) | -0.021<br>(0.059) | 0.016<br>(0.055) | -0.089*<br>(0.049) | -0.041<br>(0.059) | -0.033<br>(0.058) | -0.071**<br>(0.033) | -0.015<br>(0.034) | -0.009<br>(0.047) | -0.043<br>(0.040) | 0.054<br>(0.065) | 0.136**<br>(0.059) |
| 6 | 0.251**<br>(0.109) | -0.006<br>(0.092) | 0.108<br>(0.087) | -0.065<br>(0.080) | -0.062<br>(0.112) | -0.157*<br>(0.089) | -0.028<br>(0.044) | -0.104**<br>(0.052) | -0.194**<br>(0.086) | -0.196**<br>(0.077) | 0.060<br>(0.103) | 0.198**<br>(0.094) |
| 7 | 0.387*<br>(0.209) | 0.354*<br>(0.190) | 0.232<br>(0.297) | 0.207<br>(0.158) | 0.113<br>(0.219) | 0.032<br>(0.215) | -0.200*<br>(0.119) | 0.080<br>(0.216) | -0.199*<br>(0.112) | -0.171*<br>(0.102) | 0.608**<br>(0.263) | 0.166<br>(0.267) |
| 8 | 0.153<br>(0.152) | 0.107<br>(0.166) | 0.042<br>(0.126) | 0.225<br>(0.154) | -0.245<br>(0.190) | -0.259<br>(0.172) | 0.044<br>(0.099) | -0.085<br>(0.132) | -0.330***<br>(0.105) | 0.010<br>(0.135) | -0.089<br>(0.213) | 0.386*<br>(0.229) |
| 9 | 0.210<br>(0.443) | -0.074<br>(0.239) | 0.236<br>(0.267) | 0.421*<br>(0.251) | -0.357<br>(0.401) | -0.770**<br>(0.348) | 0.567**<br>(0.289) | 0.158<br>(0.215) | -0.260<br>(0.166) | -0.038<br>(0.094) | -0.473<br>(0.363) | -0.217<br>(0.231) |
| **No of children** |  |  |  |  |  |  |  |  |  |  |  |  |
| 0 (base) | 0.000<br>(.) | 0.000<br>(.) | 0.000<br>(.) | 0.000<br>(.) | 0.000<br>(.) | 0.000<br>(.) | 0.000<br>(.) | 0.000<br>(.) | 0.000<br>(.) | 0.000<br>(.) | 0.000<br>(.) | 0.000<br>(.) |
| 1 | -0.074**<br>(0.037) | 0.030<br>(0.034) | -0.095***<br>(0.026) | 0.001<br>(0.026) | -0.001<br>(0.033) | -0.034<br>(0.036) | 0.039**<br>(0.016) | 0.040**<br>(0.020) | -0.036<br>(0.025) | -0.032<br>(0.022) | -0.110***<br>(0.037) | -0.120***<br>(0.037) |
| 2 | -0.089*<br>(0.051) | -0.042<br>(0.045) | -0.165***<br>(0.033) | -0.091***<br>(0.034) | -0.049<br>(0.042) | 0.019<br>(0.054) | 0.008<br>(0.022) | 0.021<br>(0.026) | 0.025<br>(0.030) | 0.075**<br>(0.029) | -0.169***<br>(0.046) | -0.200***<br>(0.045) |
| 3 | 0.026<br>(0.075) | -0.049<br>(0.060) | -0.050<br>(0.060) | -0.014<br>(0.046) | 0.074<br>(0.080) | -0.022<br>(0.064) | 0.023<br>(0.032) | 0.068*<br>(0.038) | 0.090*<br>(0.047) | 0.132***<br>(0.044) | -0.120*<br>(0.066) | -0.279***<br>(0.068) |
| 4 | -0.035<br>(0.109) | -0.103<br>(0.084) | -0.124<br>(0.092) | 0.028<br>(0.068) | -0.098<br>(0.115) | -0.022<br>(0.093) | 0.111*<br>(0.065) | 0.050<br>(0.055) | 0.146<br>(0.098) | 0.060<br>(0.072) | -0.099<br>(0.120) | -0.178*<br>(0.103) |
| 5 | -0.355**<br>(0.160) | -0.135<br>(0.146) | -0.248*<br>(0.142) | -0.015<br>(0.152) | -0.116<br>(0.194) | 0.223<br>(0.203) | 0.441**<br>(0.185) | 0.133<br>(0.112) | 0.183<br>(0.125) | 0.222<br>(0.135) | -0.526**<br>(0.222) | -0.148<br>(0.184) |
| 6 | -0.246<br>(0.344) | -0.105<br>(0.186) | -0.126<br>(0.213) | 0.009<br>(0.208) | 0.187<br>(0.309) | 0.295<br>(0.334) | 0.279<br>(0.248) | 0.217<br>(0.195) | -0.102<br>(0.210) | 0.025<br>(0.139) | 0.627<br>(0.575) | -0.474<br>(0.346) |
| 7 | -0.262<br>(0.699) | 0.212<br>(0.412) | 0.303<br>(0.556) | 0.076<br>(0.317) | 0.407<br>(0.618) | 1.141***<br>(0.406) | 1.116***<br>(0.292) | -0.106<br>(0.280) | 0.187<br>(0.129) | 0.334<br>(0.375) | -0.194<br>(0.760) | -0.067<br>(0.463) |
| 8 |  |  |  |  |  |  |  |  |  |  |  |  |
| **Mode of Interview** |  |  |  |  |  |  |  |  |  |  |  |  |
| Face to Face & CAPI (base) | 0.000<br>(.) | 0.000<br>(.) | 0.000<br>(.) | 0.000<br>(.) | 0.000<br>(.) | 0.000<br>(.) | 0.000<br>(.) | 0.000<br>(.) | 0.000<br>(.) | 0.000<br>(.) | 0.000<br>(.) | 0.000<br>(.) |
| Telephone & CAPI | -0.016<br>(0.039) | -0.070*<br>(0.037) | -0.006<br>(0.037) | -0.060**<br>(0.029) | -0.003<br>(0.044) | -0.016<br>(0.038) | 0.002<br>(0.021) | -0.003<br>(0.021) | 0.018<br>(0.031) | 0.032<br>(0.030) | 0.092*<br>(0.048) | -0.028<br>(0.038) |
| Face to Face & Hard copy | 0.071<br>(0.305) | 0.043<br>(0.214) | 0.092<br>(0.241) | -0.355**<br>(0.148) | -0.144<br>(0.102) | -0.158<br>(0.234) | 0.131*<br>(0.068) | -0.008<br>(0.060) | -0.218<br>(0.228) | -0.014<br>(0.105) | 0.087<br>(0.278) | 0.109<br>(0.359) |
| Telephone & Hard copy | -0.730<br>(0.785) | 0.430***<br>(0.130) | -0.309<br>(0.332) | 0.296<br>(0.193) | 0.453**<br>(0.178) | 0.021<br>(0.151) | 0.128<br>(0.101) | 0.214<br>(0.173) | -0.062<br>(0.072) | 0.027<br>(0.087) | 0.527**<br>(0.237) | -0.181<br>(0.285) |
| Constant | -0.135<br>(0.133) | -0.091<br>(0.111) | 0.109<br>(0.097) | 0.051<br>(0.093) | 0.191<br>(0.120) | 0.085<br>(0.107) | 0.132**<br>(0.060) | 0.075<br>(0.063) | 0.229**<br>(0.096) | -0.269***<br>(0.088) | -0.082<br>(0.127) | -0.086<br>(0.113) |
| Observations | 24357 | 27854 | 24192 | 27622 | 24336 | 27825 | 23622 | 26566 | 21731 | 23322 | 24355 | 27848 |
| Groups | 5937 | 6149 | 5926 | 6131 | 5945 | 6149 | 5883 | 6060 | 5806 | 5902 | 5936 | 6153 |
| R-squared (within) | 0.679 | 0.672 | 0.781 | 0.782 | 0.648 | 0.664 | 0.891 | 0.899 | 0.811 | 0.804 | 0.590 | 0.598 |

Note: Each model allows for individual fixed effects (suppressed), and clusters standard errors at the household level to account for the fact that all individuals are interviewed in a household. All outcome variables are standardised to mean 0 and standard deviation 1. Standard errors in parentheses. Significance levels\* p<0.10 \*\* p<0.05 \*\*\* p<0.01

**Table S4.**
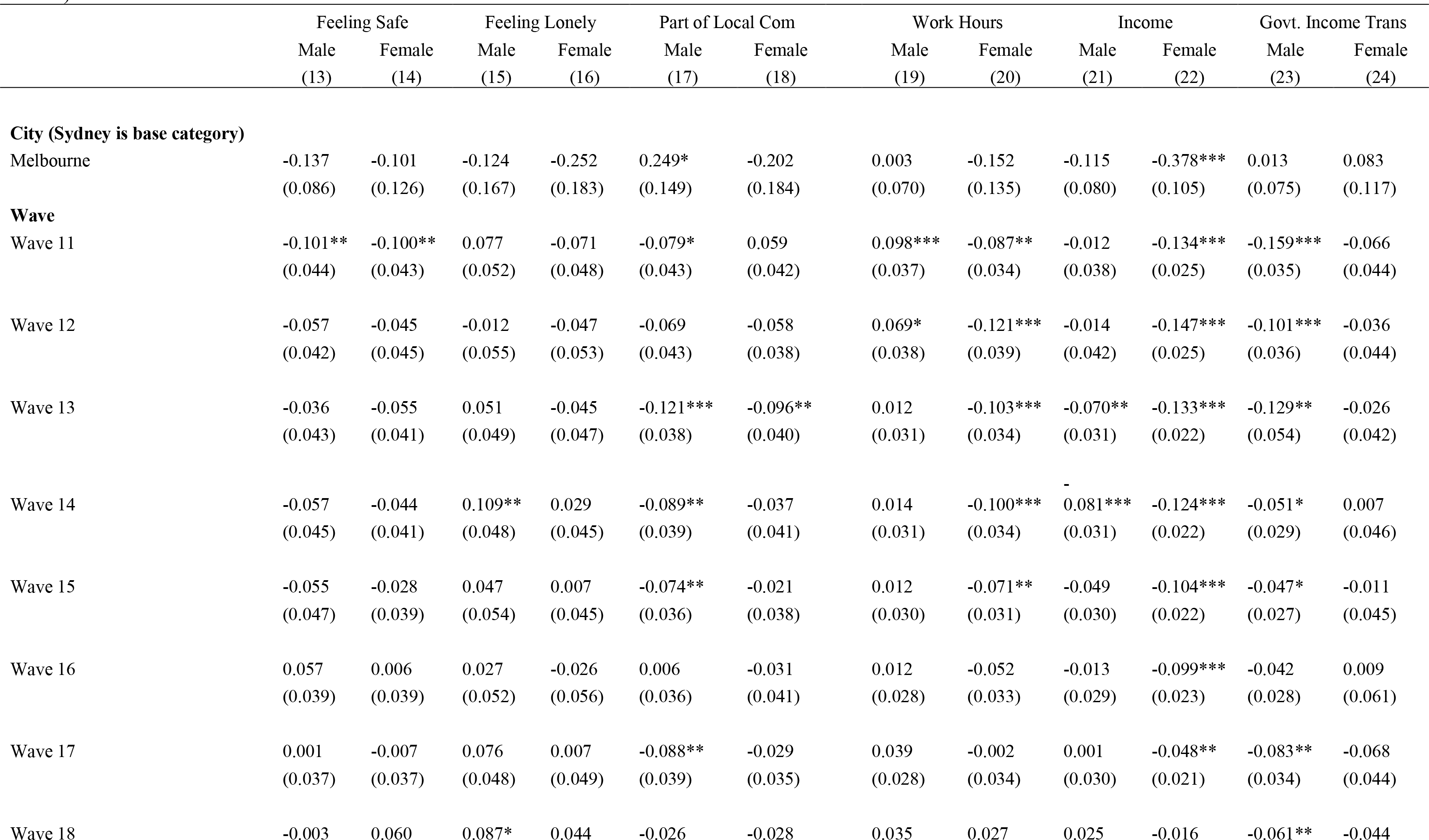
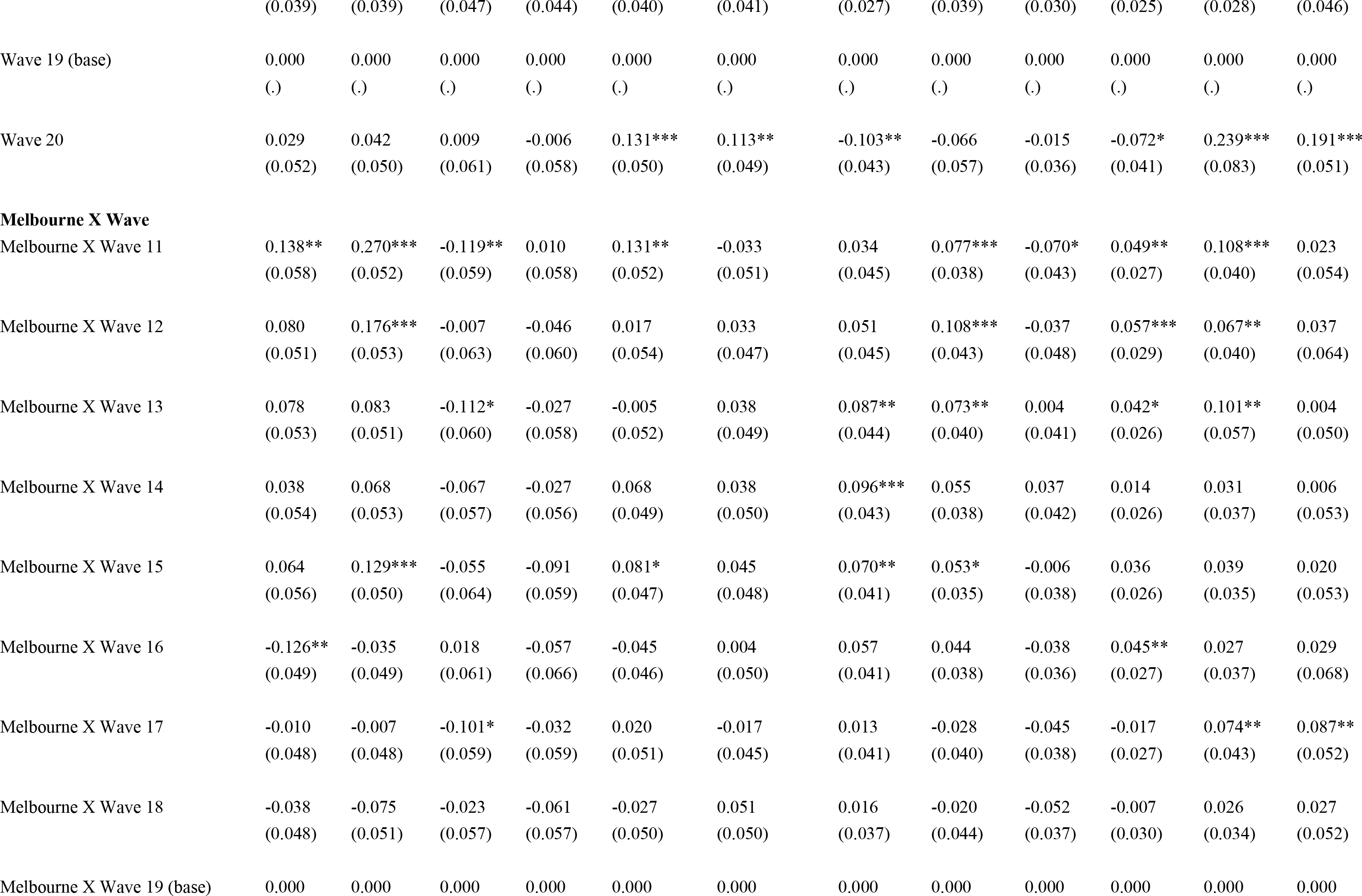
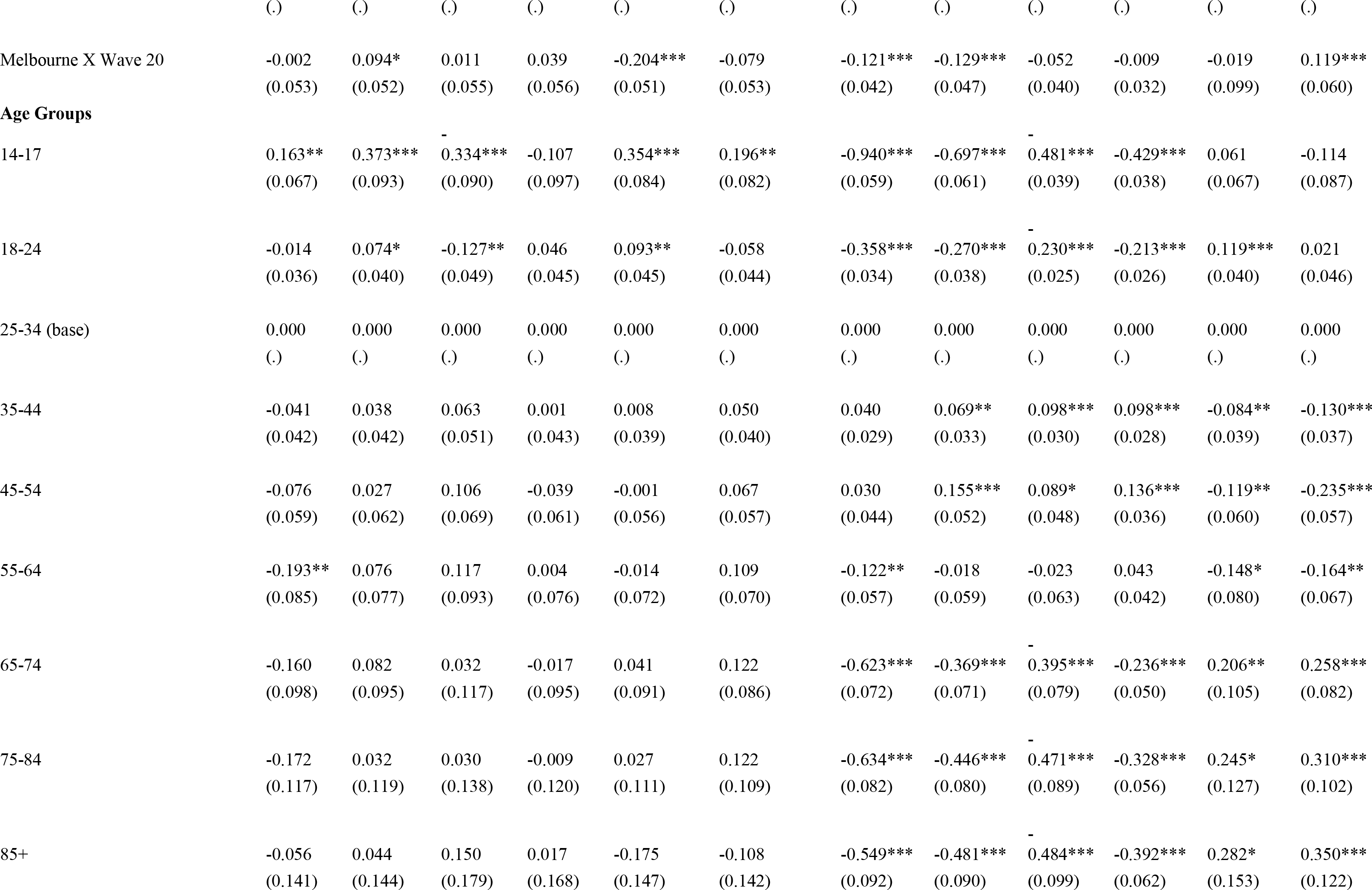
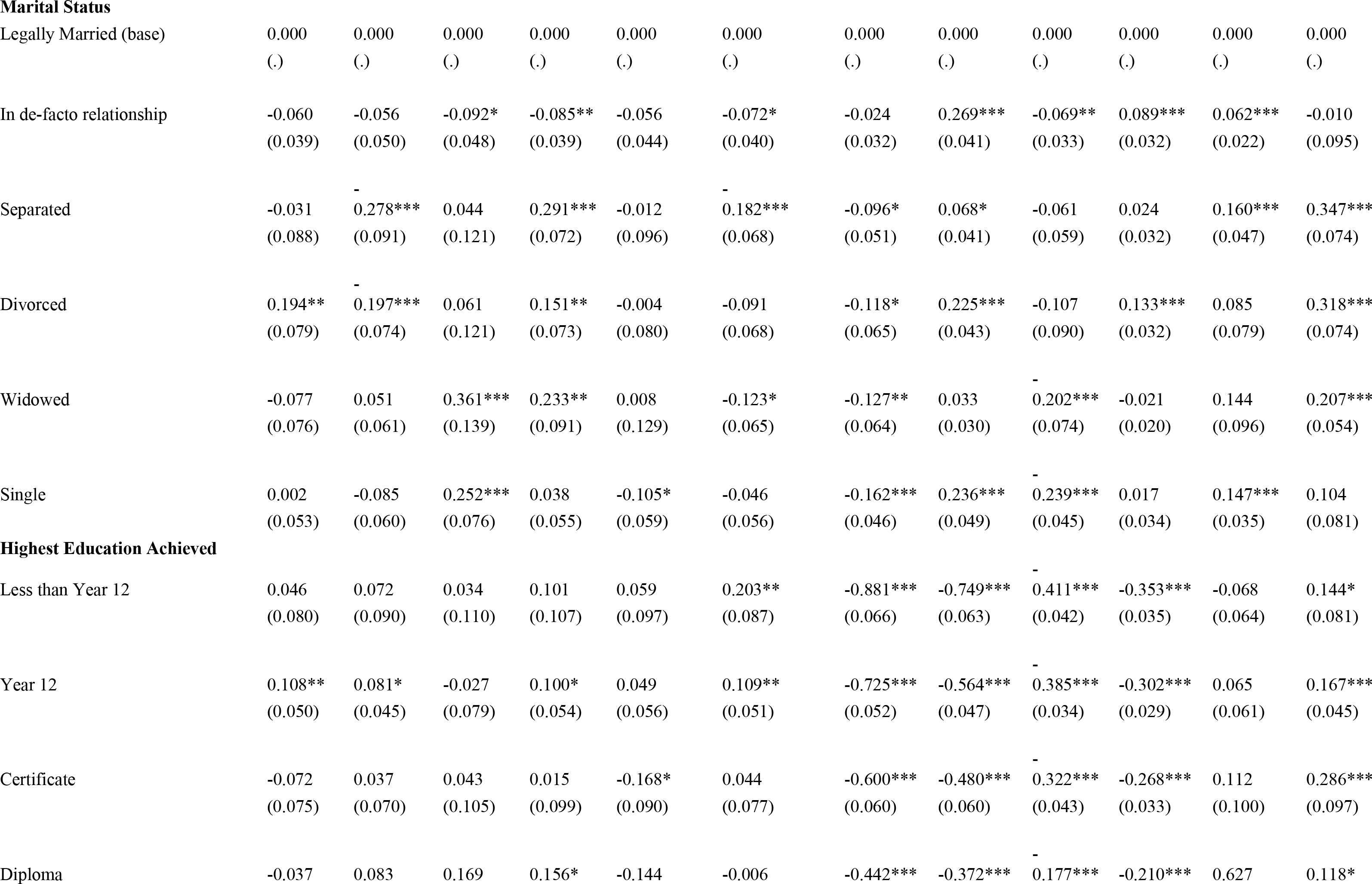
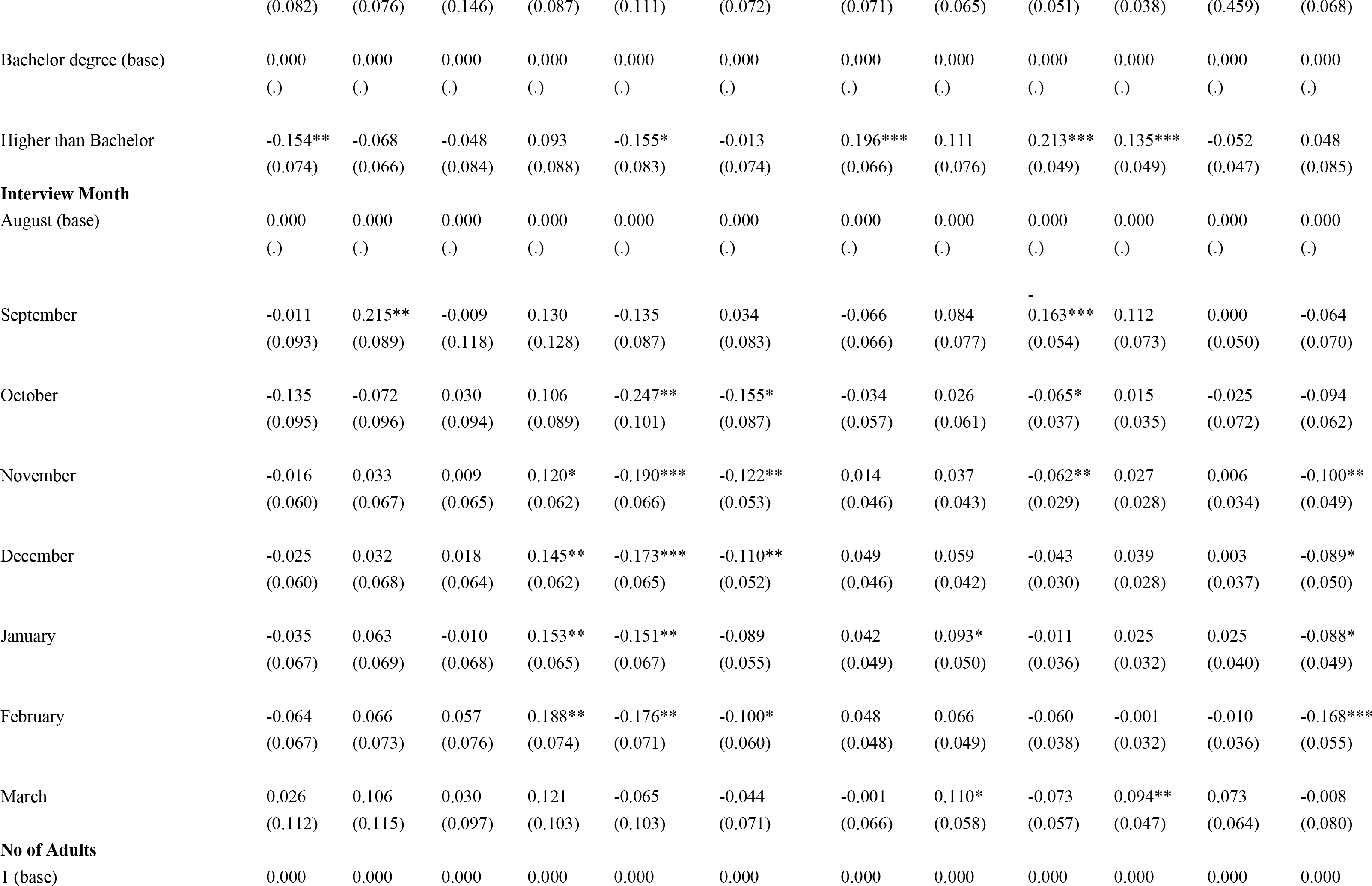
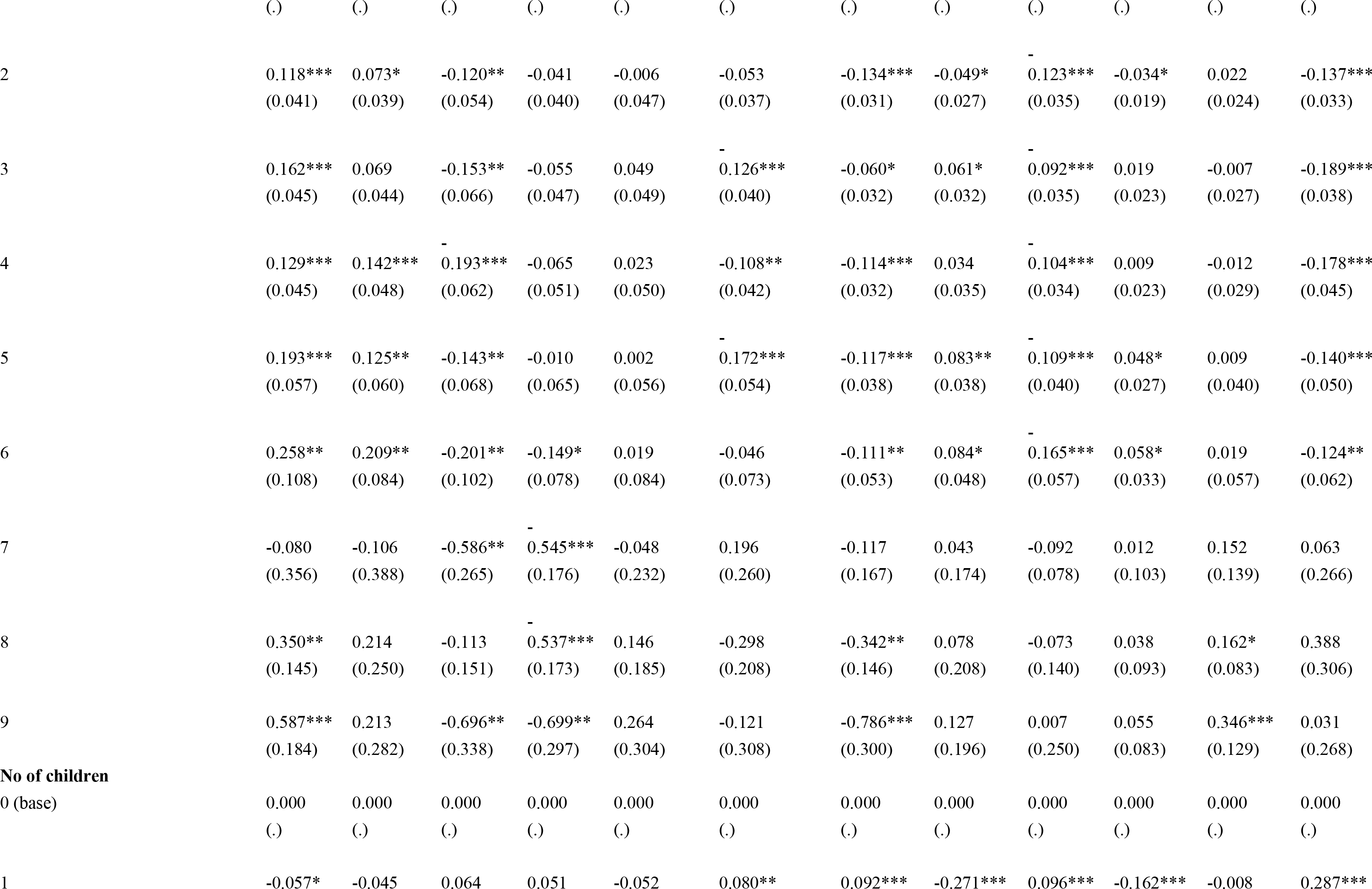
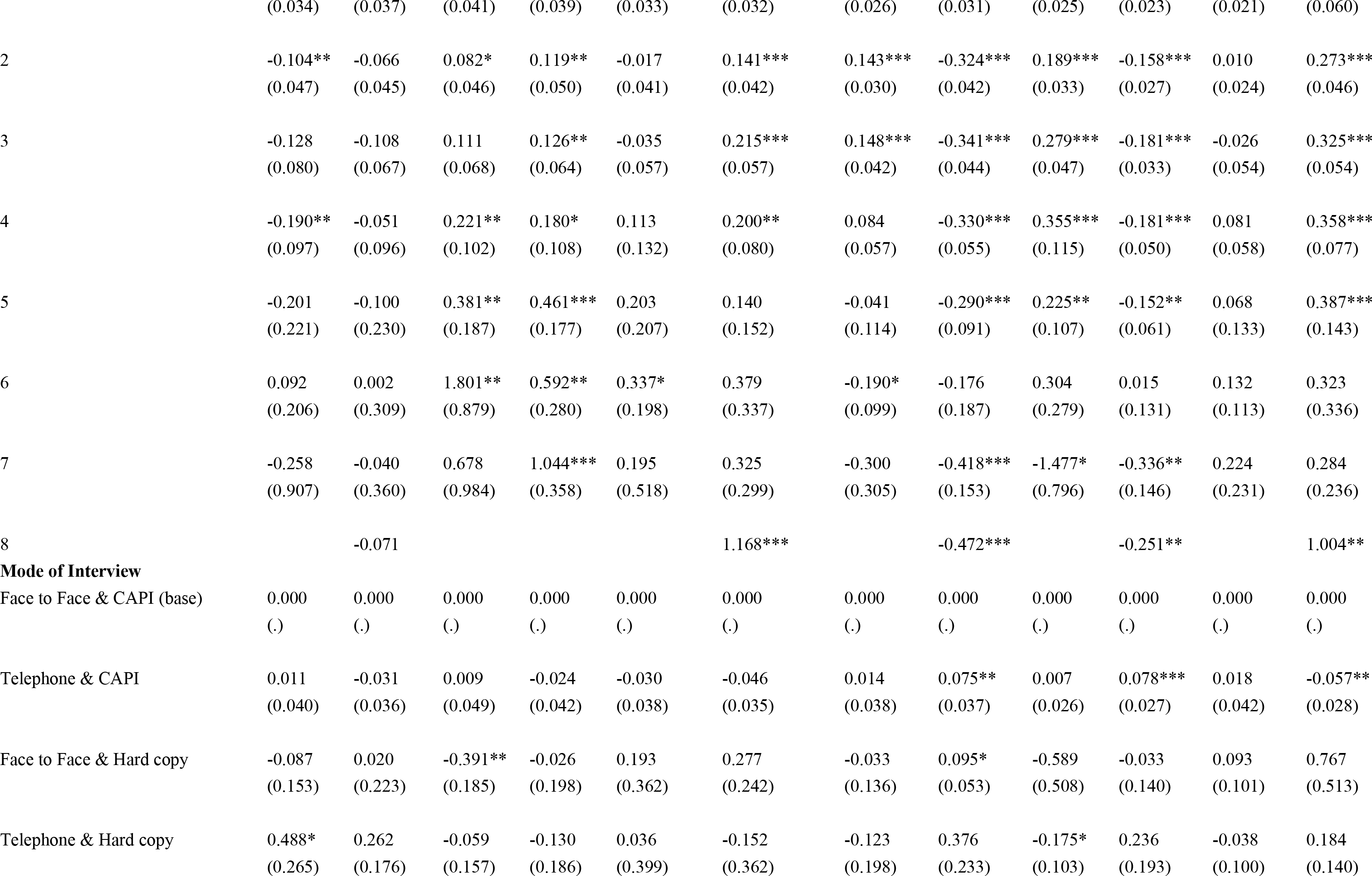
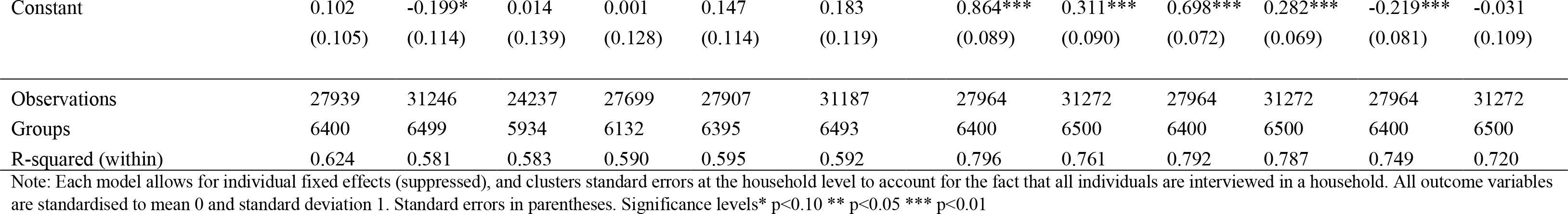
Full estimation results for social connectedness and labor supply and income (estimated treatment effects expressed in standardised to mean 0, std. dev. 1)

**Table S5.** Test of equality of coefficient between men and women with children

| | Mother × Melbourne<br>× Wave20 (std. dev.) | Standard Err. | P-Value |
| --- | --- | --- | --- |
|  | (1) | (2) | (3) |
| [1] Health domain |  |  |  |
| Mental health | -0.120 | 0.152 | 0.428 |
| General health | -0.189 | 0.123 | 0.124 |
| Bodily pain | 0.156 | 0.165 | 0.341 |
| [2] Health behaviors domain |  |  |  |
| BMI | 0.026 | 0.076 | 0.735 |
| Frequency of Alcohol<br>Consumptions | -0.223 | 0.126 | 0.075 |
| Frequency of Physical Activity | 0.354 | 0.171 | 0.038 |
| [3] Social connectedness domain |  |  |  |
| Feeling safe | 0.283 | 0.132 | 0.032 |
| Feeling lonely | 0.154 | 0.166 | 0.353 |
| Feeling part of local community | 0.086 | 0.13 | 0.506 |
| [4] Labor supply & income domain |  |  |  |
| Weekly Work Hours | -0.174 | 0.121 | 0.150 |
| Weekly Income from all Sources | 0.035 | 0.107 | 0.744 |
| Government Income Support | 0.080 | 0.108 | 0.457 |

Note: This tables reports estimates from a sample of individuals with young children. In column (1) we reported the interaction coefficient of treatment effect with mothers, expressed in standard deviations and in column (2) we reported the standard error of the interaction effect. In column (3) we reported are p-values of a test of equality of estimated coefficients between women and men with children, namely that lockdown effects were the same for fathers and mothers. The estimates are obtained from a difference-in-difference model in which we regressed a specific outcome on a treatment group indicator of having been located in Melbourne during lockdown in 2020. Treatment effects are interpreted as outcome change in Melbourne (2019-2020) relative to outcome change in Sydney (2019-2020), holding constant individual-specific effects, city-specific mean differences (base: Sydney), time trends (wave indicators), and city-specific time trends (specified as interactions between city indicator and wave indicator). Each regression applies sample weights to make the sample representative of the population, whereby sample weights are chosen either for the person or the self-completion questionnaire, depending on the outcome.

**Table S6.** P-values of test that lockdown effect was the same at the beginning and the end of lockdown

|  | Women | Men |
| --- | --- | --- |
|  | (1) | (2) |
| Mental health: with children | 0.109 | 0.656 |
| Mental health: without children | 0.528 | 0.136 |
| General health: with children | 0.030 | 0.559 |
| General health: without children | 0.706 | 0.049 |
| BMI: with children | 0.282 | 0.045 |
| BMI: without children | 0.366 | 0.095 |
| Alcohol frequency: with children | 0.749 | 0.661 |
| Alcohol frequency: without children | 0.238 | 0.674 |
| Physical activity: with children | 0.680 | 0.258 |
| Physical activity: without children | 0.601 | 0.027 |
| Perceptions of safety: with children | 0.836 | 0.782 |
| Perceptions of safety: without children | 0.734 | 0.858 |
| Feeling lonely: with children | 0.882 | 0.186 |
| Feeling lonely: without children | 0.830 | 0.058 |
| Work hours: with children | 0.023 | 0.033 |
| Work hours: without children | 0.641 | 0.971 |
| Government transfers: with children | 0.352 | 0.109 |
| Government transfers: without children | 0.107 | 0.048 |

Note: reported are p-values of a test of equality of estimated coefficients, namely that lockdown effects were different between the beginning (<40 days) and the end of lockdown (71-112 days). The estimates are obtained from a difference-in-difference model in which we regressed a specific outcome on a treatment group indicator, where treatment group indicators are separated by days since lockdown started (<40days, 40-70 days, 71-112 days, outside of lockdown (113+ days). Treatment effects are interpreted as outcome change in Melbourne (2019-2020) relative to outcome change in Sydney (2019-2020), holding constant individual-specific effects, city-specific mean differences (base: Sydney), time trends (wave indicators), and city-specific time trends (specified as interactions between city indicator and wave indicator). Each regression applies sample weights to make the sample representative of the population, whereby sample weights are chosen either for the person or the self-completion questionnaire, depending on the outcome.

**Table S7.** Differences in COVID-19 exposure, risk perceptions and income effects of first lockdown between Melbourne and Sydney

|  | Male<br>(1) | Female<br>(2) |
| --- | --- | --- |
| *Panel A: Have you had COVID-19? [No: 0, Yes:1]* |  |  |
| Lockdown | 0.001<br>(0.002) | -0.002<br>(0.003) |
| Observations | 3028 | 2685 |
| Mean | .0036 | .0037 |
| Implied % change | 27.7 | 54.4 |
| *Panel B: What is your risk of getting infected with COVID-19? [0-100%]* |  |  |
| Lockdown | 3.760***<br>(0.920) | 2.096**<br>(0.930) |
| Observations | 2859 | 2581 |
| Mean | 24.7 | 22.1 |
| Implied % change | 15.2 | 9.4 |
| *Panel C: In case you get infected, what is your risk of getting seriously ill? [0-100%]* |  |  |
| Lockdown | 0.925<br>(1.115) | -1.783<br>(1.090) |
| Observations | 2842 | 2563 |
| Mean | 37.7 | 31.9 |
| Implied % change | 2.5 | 5.6 |
| *Panel D: Last financial year income (disposable) [in Std. Dev.]* |  |  |
| Lockdown | -0.017<br>(0.043) | 0.086*<br>(0.050) |
| Observations | 24427 | 27946 |
| Std. dev. | 72555 | 57521 |
| \$ difference | -1,233 | 4,946 |

Note: *Panel A-C*: Reported are estimates of differences between Melbourne and Sydney in *COVID-19* exposure “Have had *COVID-19* already” (*Panel A*); in risk of getting infected with *COVID-19* (*Panel B*), and in your risk of getting seriously ill if infected (*Panel C*). Models were estimated on sample of participants who were asked this question in the 2020 HILDA *COVID-19* module. Each model controls flexibly for age group indicators, current marital status, higher level of education achieved, month of interview and interview modus (telephone vs in-person), number of dependent children in household, number of adult household members, country of birth. *Panel D*: the outcome is last financial year total income from all sources, whereby the financial year refers to July -June of any given year, and thus covers any income effects through the first national lockdown. The estimation model in *Panel D* is a standard difference-in-difference model as in our benchmark models, see notes to Figure 2 for model specification. Standard errors in parentheses \* $p < .10$ , \*\* $p < 0.05$ , \*\*\* $p < 0.01$

**Figure S1.**
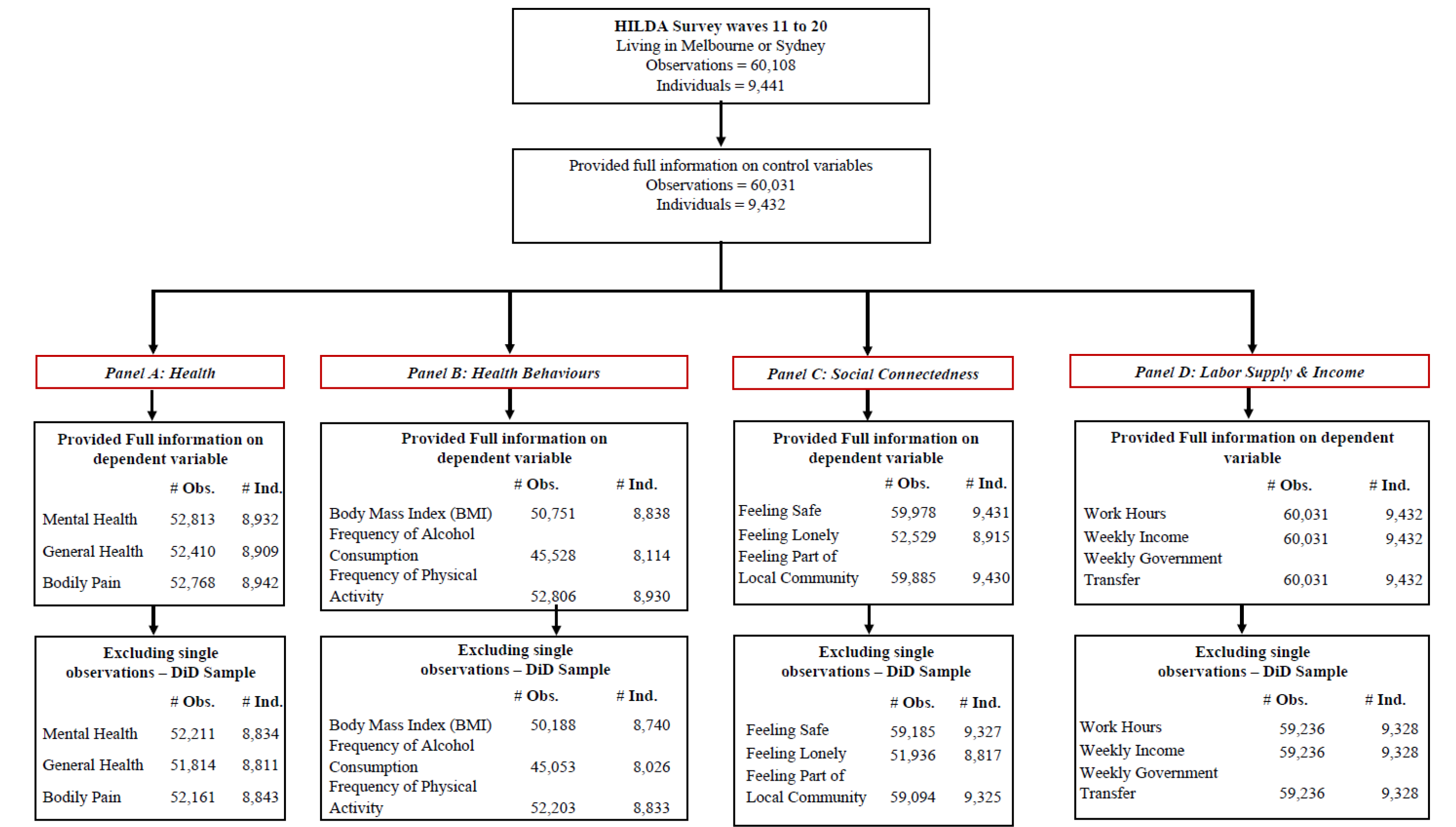
Evolution of sample size from original data to estimation sample

**Figure S2.**
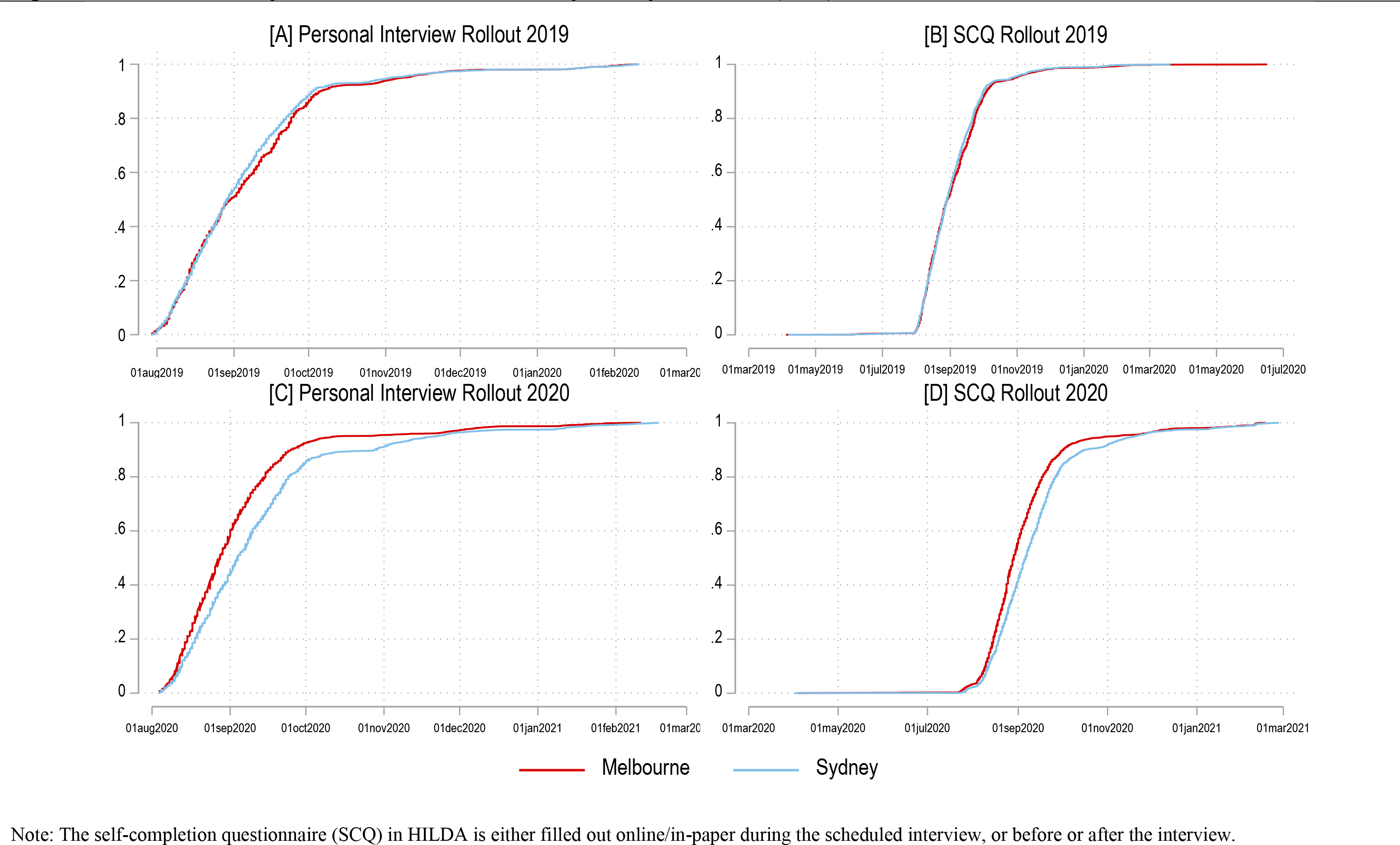
Rollout dates of personal interview and self-completion questionnaire (SCQ), 2019 versus 2020

**Figure S3.**
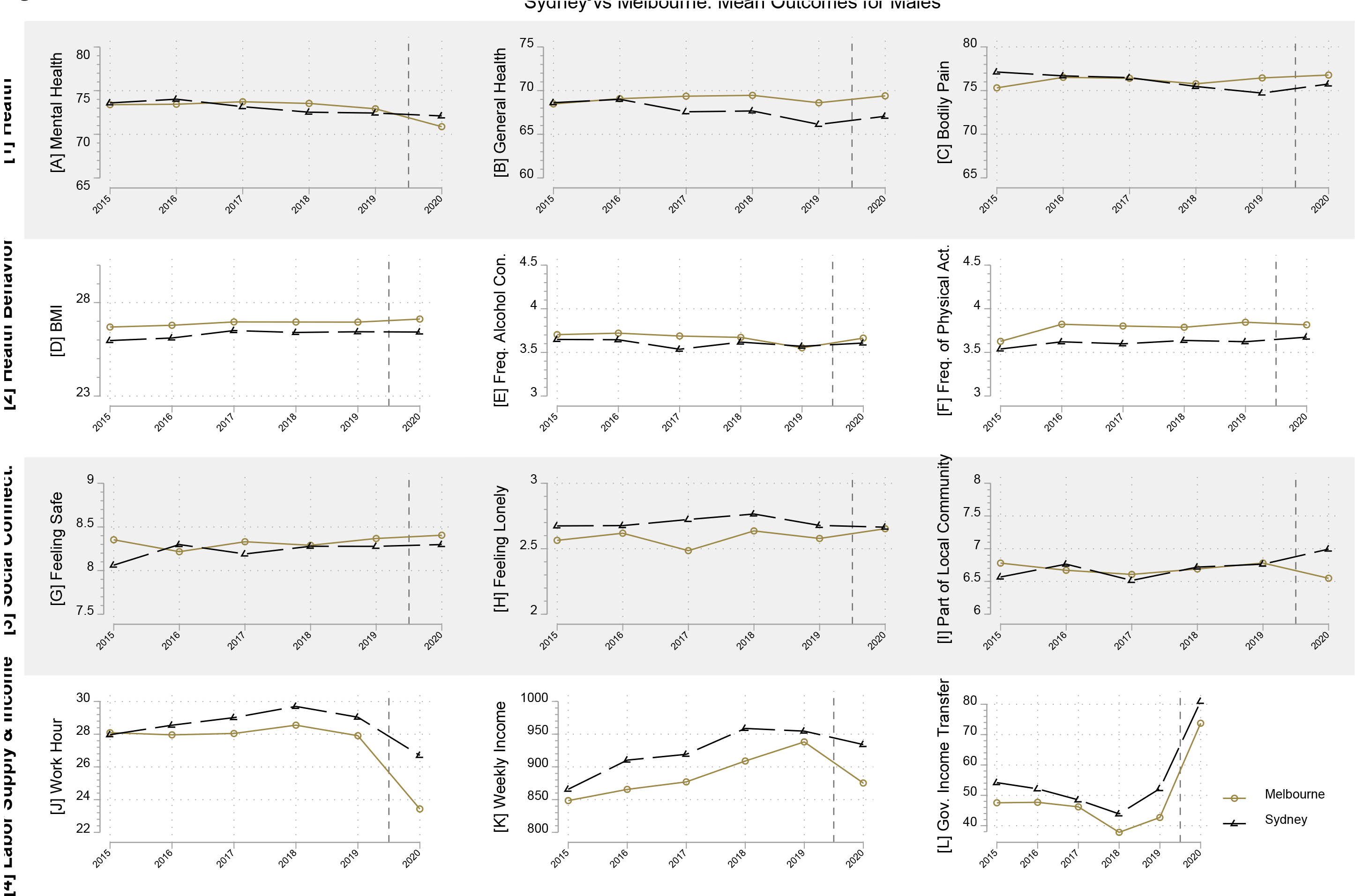
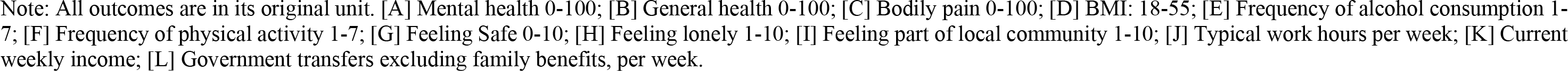
Time trends in outcomes between Melbourne and Sydney, male sample

**Figure S4.**
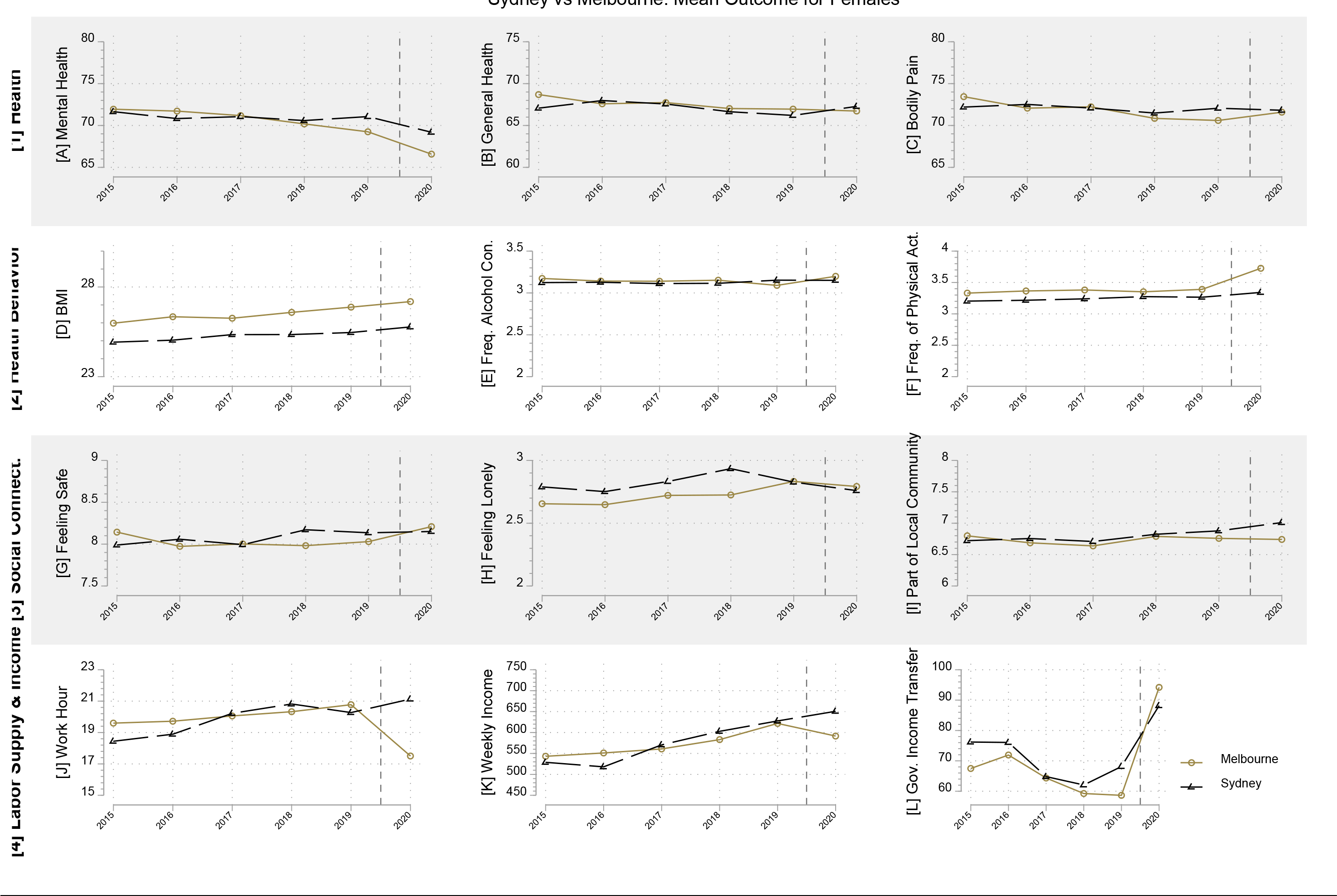
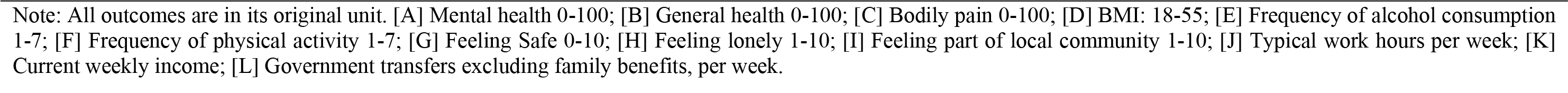
Time trends in outcomes between Melbourne and Sydney, female sample

**Figure S5.**
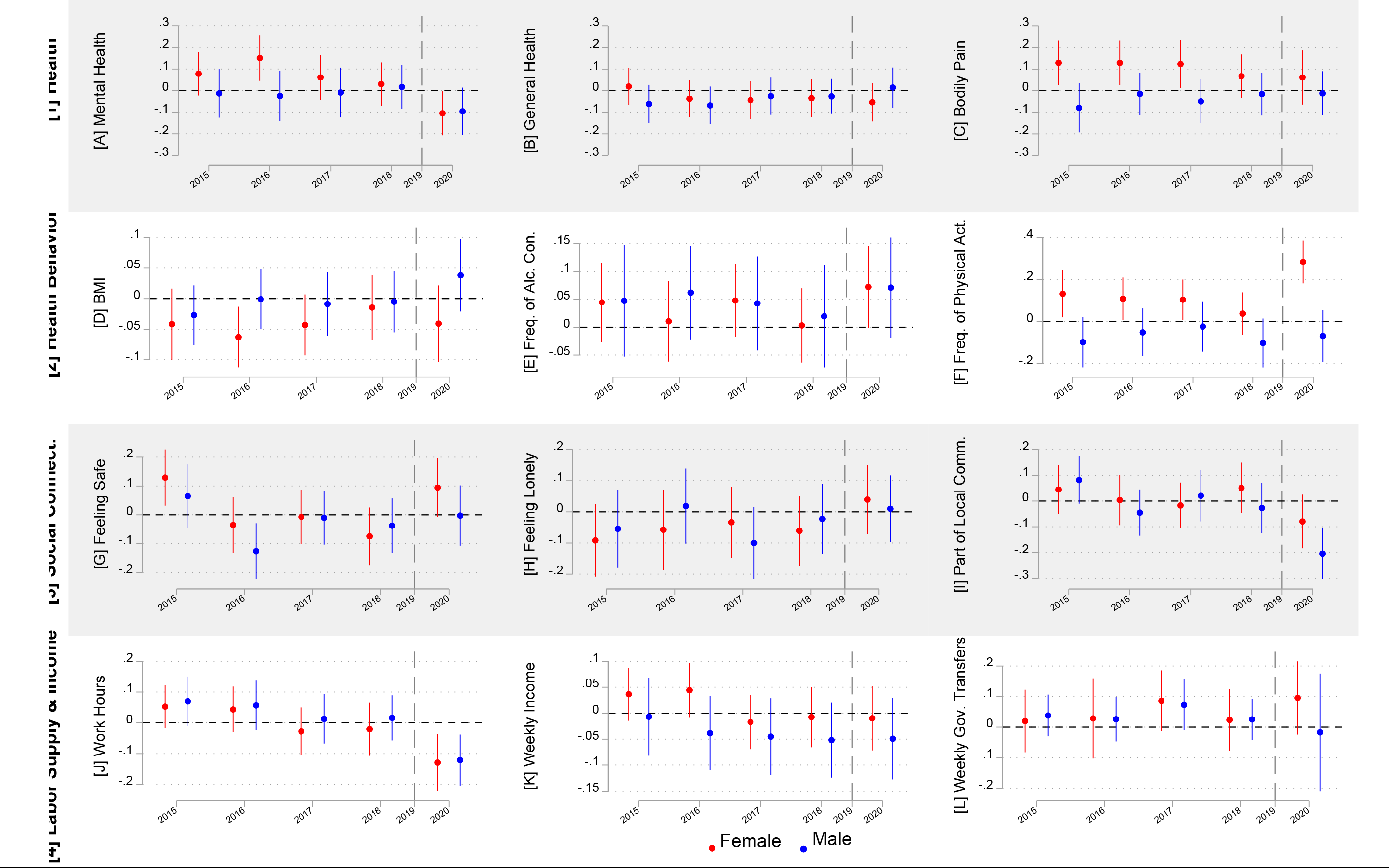
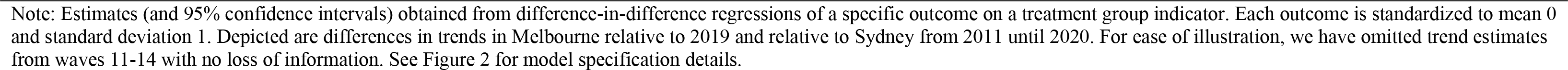
Test of the common trend assumption necessary for reliable difference-in-difference estimates of the treatment effect (expressed in std. dev.), by sex

**Figure S6.**
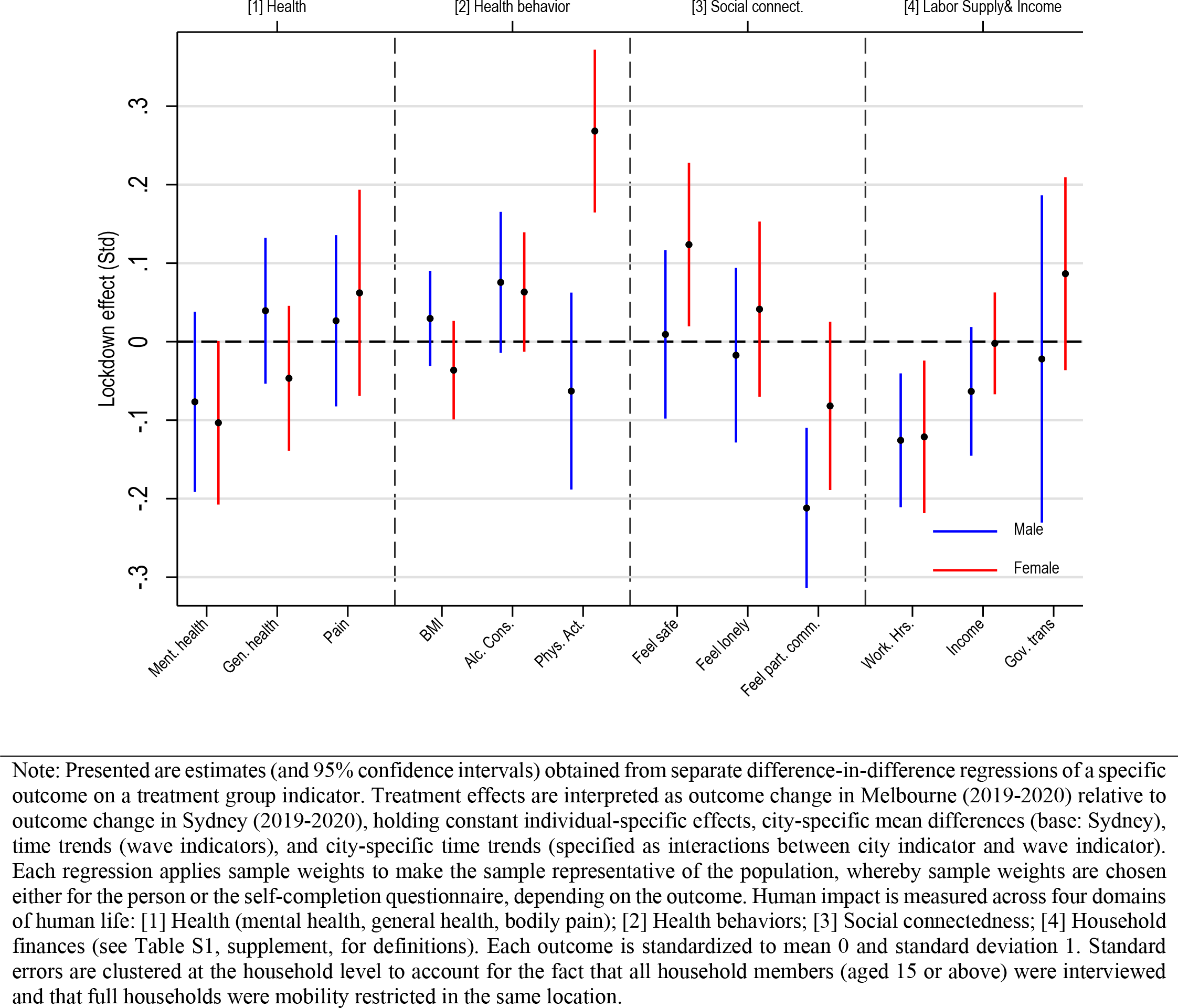
Robustness check: Drop individuals interviewed outside the lockdown period (N=267)

**Figure S7.**
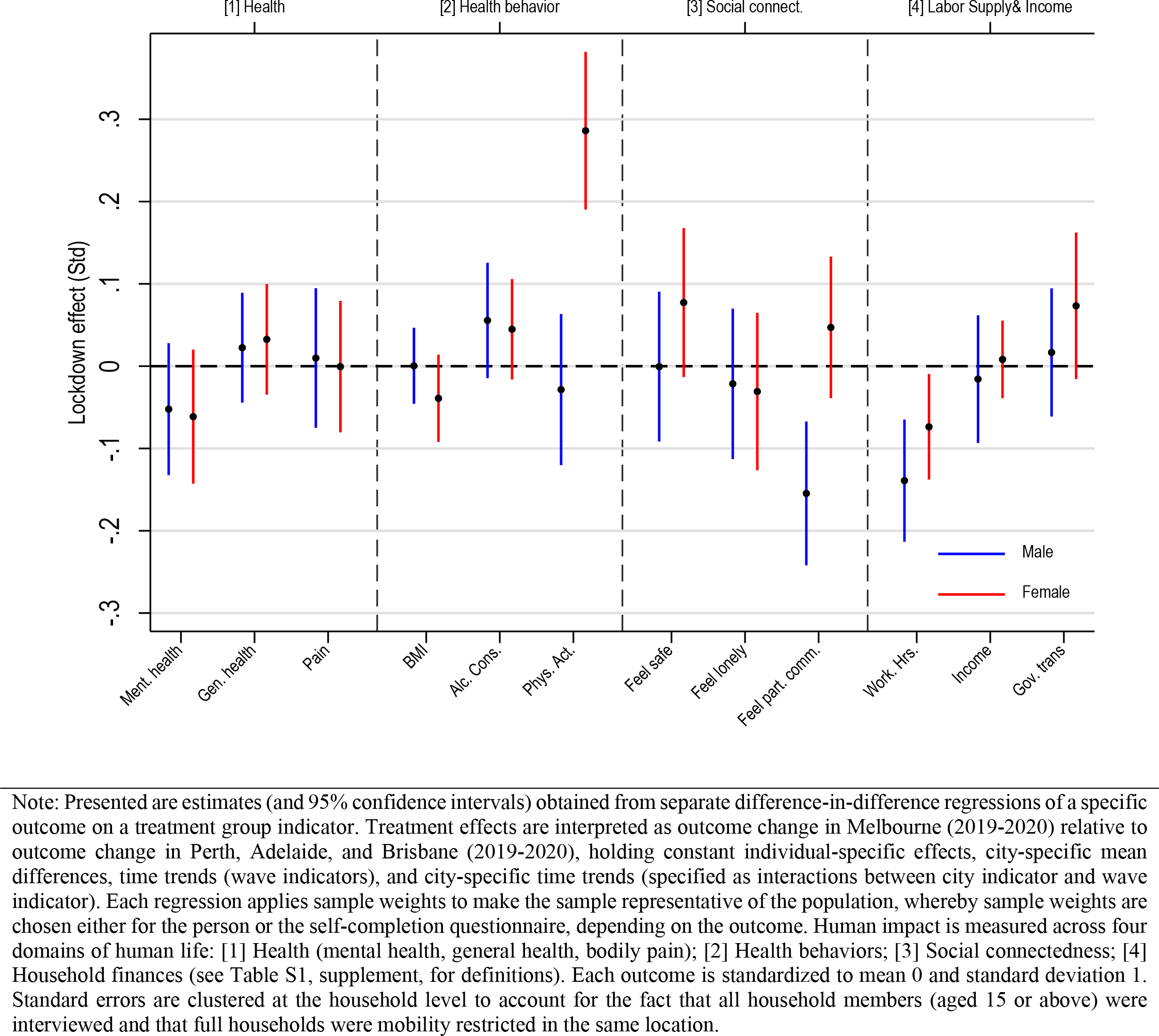
Robustness check: Use as control group Brisbane, Perth and Adelaide instead of Sydney

**Figure S8.**
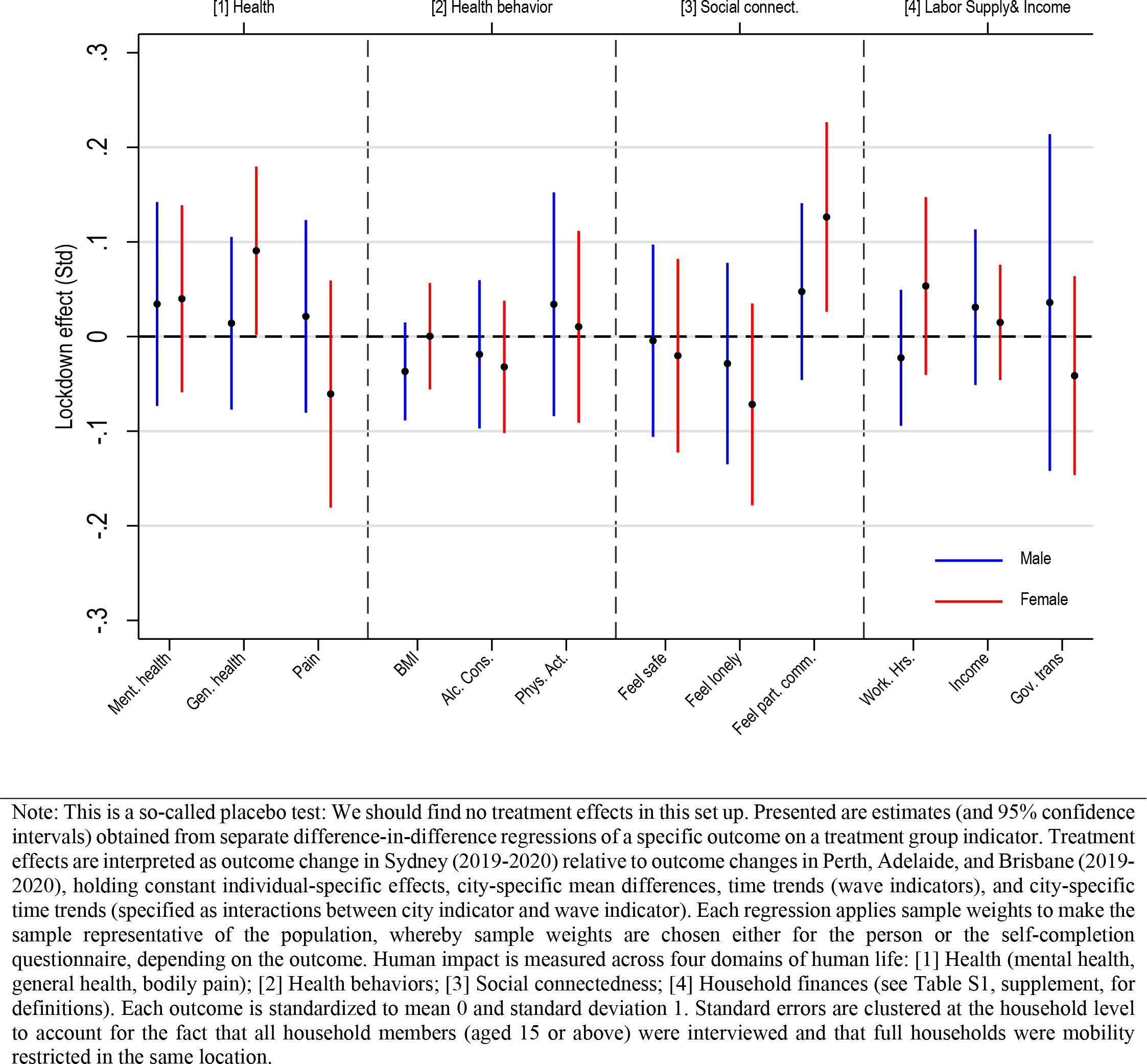
Robustness check: Placebo treatment – use Sydney as treatment group and Perth, Adelaide, and Brisbane as control groups

**Figure S9.**
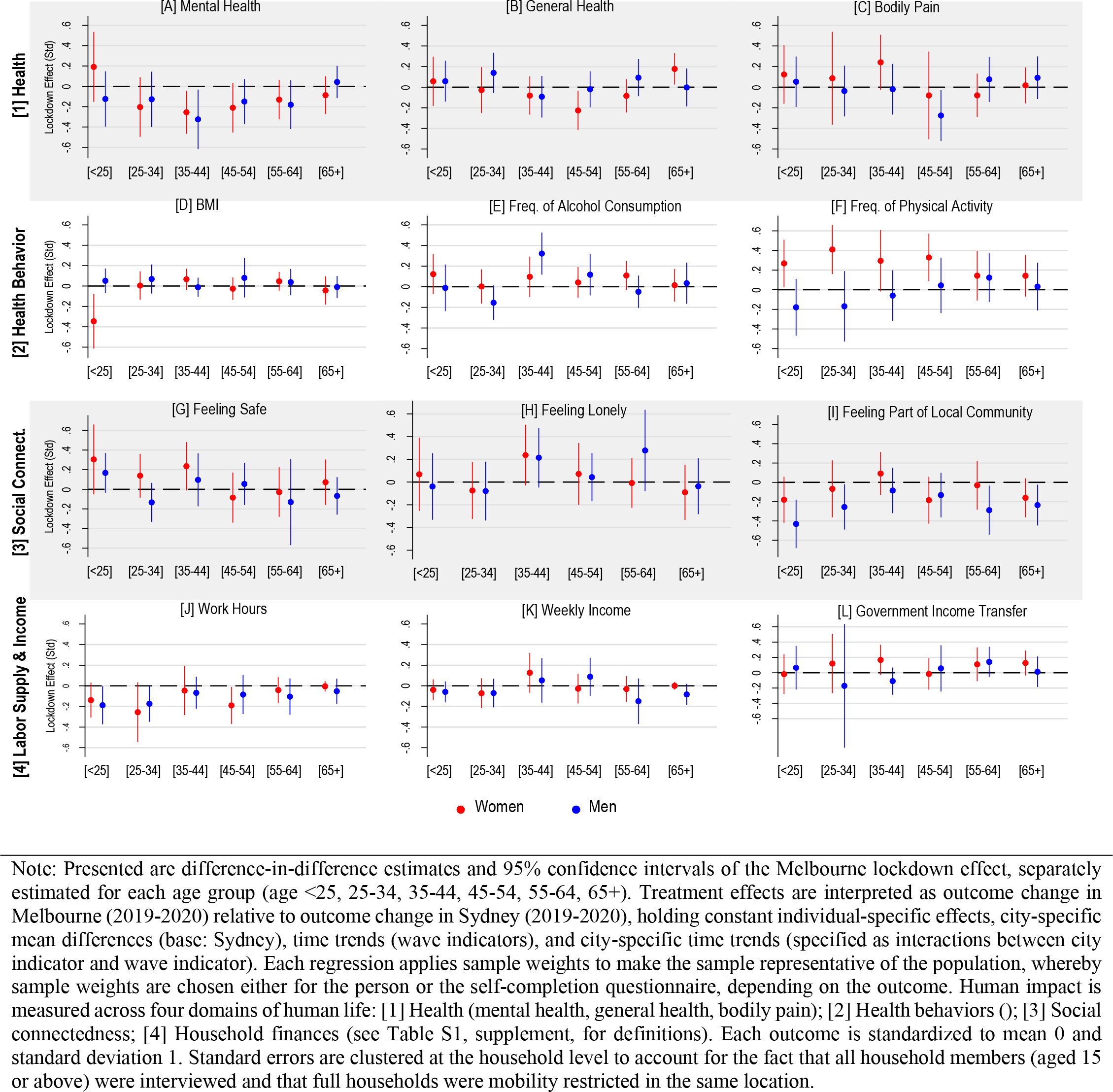
Lockdown treatment effects (in std. dev.), by age groups

**Figure S10.**
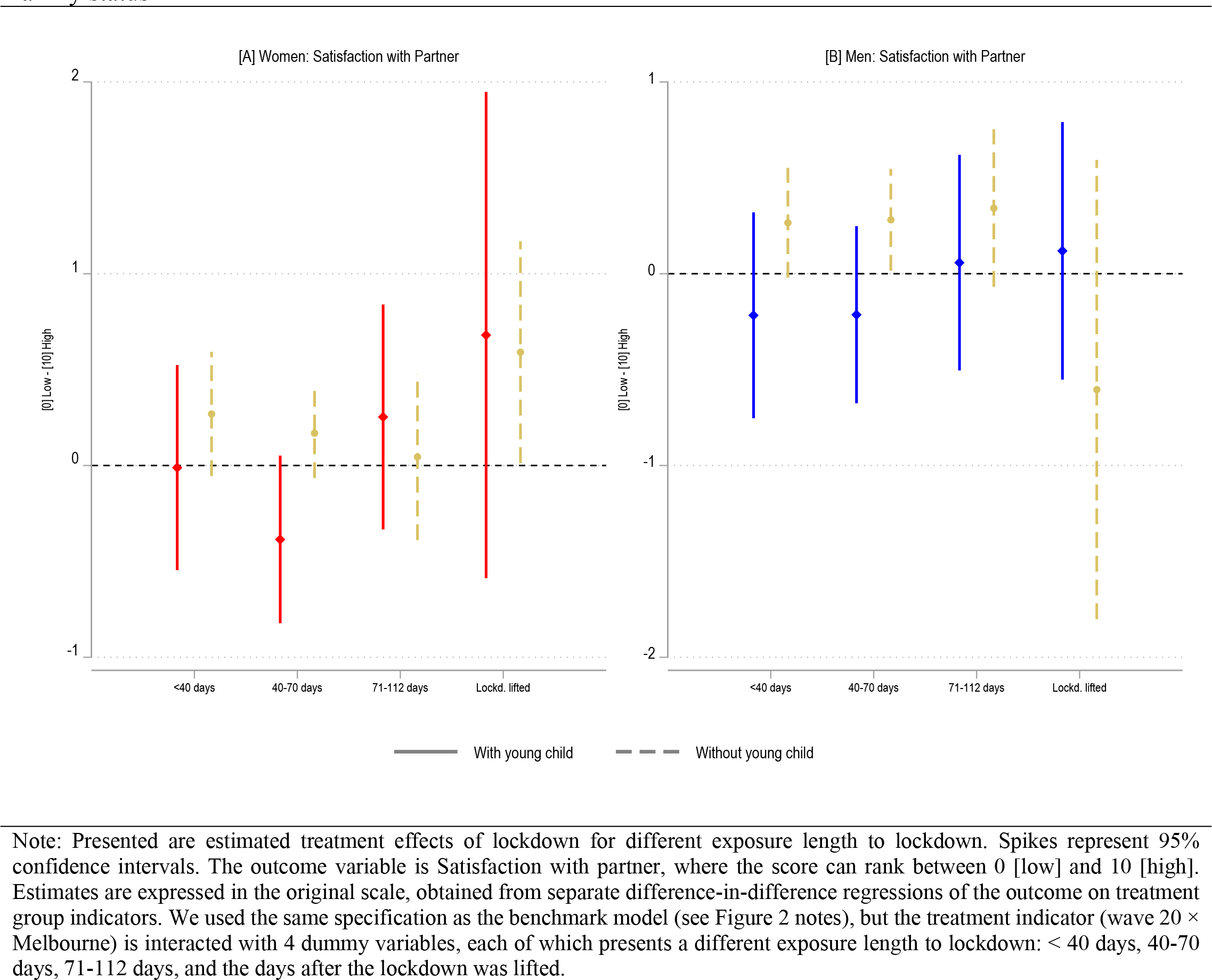
Lockdown treatment effect on satisfaction with partners (original scale), by length of exposure andfamily status

## Notes

### Competing Interest Statement

The authors have declared no competing interest.

### Funding Statement

This study was funded by the Australian Research Council (ARC).

### Author Declarations

We used data from the Household Income and Labour Dynamics in Australia (HILDA) survey which is publically available. This paper uses unit record data from the Household, Income and Labour Dynamics in Australia (HILDA) Survey, conducted by the Melbourne Institute of Applied Economic and Social Research on behalf of the Australian Government Department of Social Services (DSS) (Release 20, doi:10.26193/PI5LPJ, ADA Dataverse.) The findings and views reported in this paper, however, are those of the authors and should not be attributed to the Australian Government, the DSS, or the Melbourne Institute. The data used are available free of charge to researchers through the National Centre for Longitudinal Data Dataverse at the Australian Data Archive (https://dataverse.ada.edu.au/dataverse/ncld). Access is subject to approval by the Australian Government Department of Social Services and is conditional on signing a license specifying terms of use.

